# Accurate Prostate Cancer Detection based on Circulating Tumor Cell Profiling

**DOI:** 10.1101/2022.05.11.22274934

**Authors:** Sewanti Limaye, Simon Chowdhury, Nitesh Rohatgi, Anantbhushan Ranade, Nelofer Syed, Johann Riedemann, Raymond Page, Darshana Patil, Dadasaheb Akolkar, Vineet Datta, Revati Patil, Kiran Bendale, Pooja Fulmali, Pradeep Fulmali, Sachin Apurwa, Stefan Schuster, Sudha S Murthy, Chirantan Bose, Jinumary John, Ajay Srinivasan, Rajan Datar

**Author notes:** **CORRESPONDING AUTHOR** Dr. Ajay Srinivasan.

## Abstract

**Background:** Evaluation of serum prostate specific antigen (PSA) is a part of standard prostate cancer diagnostic work-up in symptomatic males as well as for elective prostate cancer screening in asymptomatic males. The low specificity of serum PSA leading to an inability to effectively differentiate prostate cancer from benign prostate conditions is a persistent clinical challenge. Further, the low sensitivity of serum PSA leading to false negatives can miss high-grade / aggressive prostate cancers.

**Objective:** We describe a non-invasive prostate cancer detection test based on functional enrichment of prostate adenocarcinoma associated circulating tumor cells (PrAD-CTCs) from blood samples and their identification via immunostaining for pan-cytokeratins (PanCK), prostate specific membrane antigen (PSMA), alpha methyl-acyl coenzyme-A racemase (AMACR), epithelial cell adhesion molecule (EpCAM) and common leucocyte antigen (CD45).

**Design, Setting, and Participants:** The analytical validation studies used VCaP reference prostate cancer cell line to evaluate the performance characteristics of the test. The clinical performance characteristics of the test were first evaluated in a case-control study with 160 known prostate cancer cases and 800 healthy males. A prospective clinical study was performed with samples from 210 suspected cases of prostate cancer.

**Outcome Measurements and Statistical Analysis:** Analytical validation established analyte stability as well as acceptable performance characteristics. The test showed 100% specificity and 100% sensitivity to differentiate prostate cancer cases from healthy individuals in the case control study and 91.2% sensitivity and 100% specificity to differentiate prostate cancers from benign prostate conditions in the prospective clinical study.

**Results and Limitations:** The test accurately detects PrAD-CTCs with high sensitivity and specificity irrespective of stage or grade (Gleason score), which translates into low risks of false negatives or overdiagnosis. The test does not detect minor non-adenocarcinoma subtypes of prostate cancer.

**Conclusions:** The high accuracy of the test offers advantages over PSA based prostate cancer detection.

## INTRODUCTION

Prostate cancer is globally the second most common malignancy and the seventh highest cause of cancer related mortality among men [1]. Detection of prostate cancer at advanced stages is associated with significant morbidity and mortality as well as reduced survival, while early-stage prostate cancer detection is associated with higher cure rate and improved survival (∼99%, 5-year [2]). At present, evaluation of serum prostate specific antigen (PSA) is part of the standard diagnostic work-up in symptomatic cases [3] but less suitable for prostate cancer screening in asymptomatic males due to low specificity [4] and significant risk of false positivity [5] which leads to overdiagnosis and overtreatment [6]. In addition, there is a risk of false negatives, especially in advanced undifferentiated prostate cancers which may have lower PSA levels [7]. More sensitive and specific methods which can provide for more effective prostate cancer detection are required to reduce morbidity and mortality from this disease [8].

Circulating tumor analytes in blood have received attention for non-radiological, non-invasive detection of prostate cancer [9]. Apart from serum tumor antigens, circulating tumor nucleic acids have been evaluated for prostate cancer detection but have reported limitations in sensitivity for localized prostate cancer [10]. Circulating tumor cells (CTCs) are viable tumor derived cells in circulation, the molecular and functional evaluation of which may be comparable to that of the tumor tissue from which they originate [11]. CTC evaluations are not prone to the limitations in sensitivity and specificity associated with circulating tumor nucleic acids or serum tumor antigens. Prior studies support the ubiquity of CTCs in prostate cancer, especially in early-stage (localized) disease; disseminated tumor cells (DTCs) released during early stages of prostate cancer are known to remain dormant in the bone marrow and result in metastatic recurrence [12]. In a study of bone marrow aspirates from 533 preoperative prostate cancer cases with localized disease (T2-4, N0), DTCs were detected in 380 cases (71.3%), irrespective of pathologic stage, Gleason grade, or PSA [13]. Another study reported CTCs in 19 (79%) of 24 treatment naïve localized prostate cancers [14]. A third study reported >90% sensitivity in 20 known prostate cancer cases and 92.6% sensitivity in 27 asymptomatic men undergoing prostate cancer screening [15]. A fourth study on pre-operative blood from 86 prostate cancer cases reported 38.4% - 62.7% CTC detection rates using CellSearch, CellCollector and EPISPOT individually, and 80.2% [16] when used together. In a fifth study, using a hybrid microfluidic-imaging along with PSA immunostaining, 38 - 222 CTCs were reported per mL in recently diagnosed cases of localized prostate cancer [17]. In a sixth study, using near-infrared dyes and EpCAM immunostaining, up to 439 CTCs per mL of blood (mean: 25 CTCs / mL; median: 10 CTCs / mL) were observed in a cohort of patients with localized prostate cancer [18]. The above studies provide evidence for the biological plausibility of CTC-based prostate cancer screening. Other studies have also shown the inability of existing technology platforms to efficiently enrich and harvest sufficient CTCs. Most prior reports on CTCs in cancer are based on epitope capture using epithelial cell adhesion molecule (EpCAM) followed by immunostaining for cytokeratins (CK). A critical limitation of this approach is its acknowledged inability to effectively enrich and detect CTCs where the expression of target biomarkers such as EpCAM and CK can be significantly lower [19–23] than tumor tissue or reference cell lines. Further, the expression of EpCAM and CK (as well as any other markers) may be even lower in CTCs undergoing epithelial to mesenchymal transition (EMT) [24].

We have previously described a functional CTC enrichment process which is immune to the limitations of epitope-based CTC enrichment and yields numerically sufficient CTCs for further applications [25]. We have also shown that CTCs thus enriched from blood of patients with prostate cancer are positive for expression of PSMA and AMACR in addition to EpCAM and PanCK as determined by fluorescence immunocytochemistry (ICC) [26]. This multi-marker CTC profiling has high specificity for adenocarcinomas (AD) which represent the vast majority (∼92%) of prostate cancers [27]. The test uses standardized fluorescence intensity (FI) thresholds for detection of marker positive cells, optimized to detect CTCs with a wide range of marker expression, especially those with significantly lower marker expression than tumor cells or reference cell lines. In this manuscript, we report the method development as well as analytical and clinical validation of this test for prostate cancer detection.

## METHODS

### Study Participants and Samples

Samples for method development, analytical validation and clinical validation studies were obtained from participants in three observational studies, TRUEBLOOD (http://ctri.nic.in/Clinicaltrials/pmaindet2.php?trialid=31879), ProState (http://ctri.nic.in/Clinicaltrials/pmaindet2.php?trialid=31713) and RESOLUTE (http://ctri.nic.in/Clinicaltrials/pmaindet2.php?trialid=30733). The TRUEBLOOD study enrolled known patients with various solid organ cancers or benign (non-malignant) conditions as well as individuals who were suspected of various cancers. The ProState study enrolled known cases of prostate cancers and symptomatic males suspected of prostate cancer. The RESOLUTE study enrolled healthy asymptomatic adults with no prior diagnosis of cancer, no current symptoms or findings suspected of cancer. All studies were approved by the Ethics Committees of the participating institutes and the sponsor and were performed in accordance with the Declaration of Helsinki. Fifteen millilitres of peripheral blood were collected from all enrolled study participants in EDTA vacutainers after obtaining written informed consent. Where possible, tissue samples were also obtained from TRUEBLOOD and ProState study participants referred for a biopsy as per Standard of Care (SoC). In addition, blood samples for research were collected, after obtaining consent, from healthy (asymptomatic) volunteers as well as recently diagnosed or suspected cancer patients who were not a part of either of the above studies but had availed of the sponsor’s services. Blood samples from suspected cases of cancers were collected prior to the patients undergoing an invasive biopsy. All biological samples were stored at 2°C - 8°C during transport to reach the clinical laboratory within 46 h. Samples used for clinical validation studies were identity masked by using blood collection vacutainers with a 10-digit alphanumeric code. Identity masking minimized potential biases resulting from differences in sample processing or interpretation of results that could have arisen due to operator’s knowledge of the sample. All samples were processed at the CAP and CLIA accredited facilities of the Study Sponsor, which also adhere to quality standards ISO 9001:2015, ISO 27001:2013 and ISO 15189:2012.

### Isolation of Primary Tumor Derived Cells

The isolation of primary tumor derived cells (TDCs) from an excised tumor (malignant / benign) was performed as described previously [25] and is also explained in **Supplementary Materials**.

### Enrichment of Circulating Tumor Cells from Peripheral Blood

Blood samples (5 mL) were processed for the enrichment of CTC from peripheral blood mononuclear cells (PBMC) as described previously [26,28]. The process is also explained in **Supplementary Materials**.

### Immunocytochemistry Profiling of CTCs

The process of ICC profiling of CTC was as described previously [26] and is also provided in **Supplementary Materials**. A schema showing the various steps of the process including CTC detection and ICC profiling is depicted in **Figure 1**. The decision matrix for assigning samples as ‘Positive’, ‘Equivocal’ or ‘Negative’ based on marker positivity and number of positive cells is provided in **Figure 2**. Samples with equivocal findings were considered positive for the purpose of prostate cancer detection by the test.

**Figure 1.**
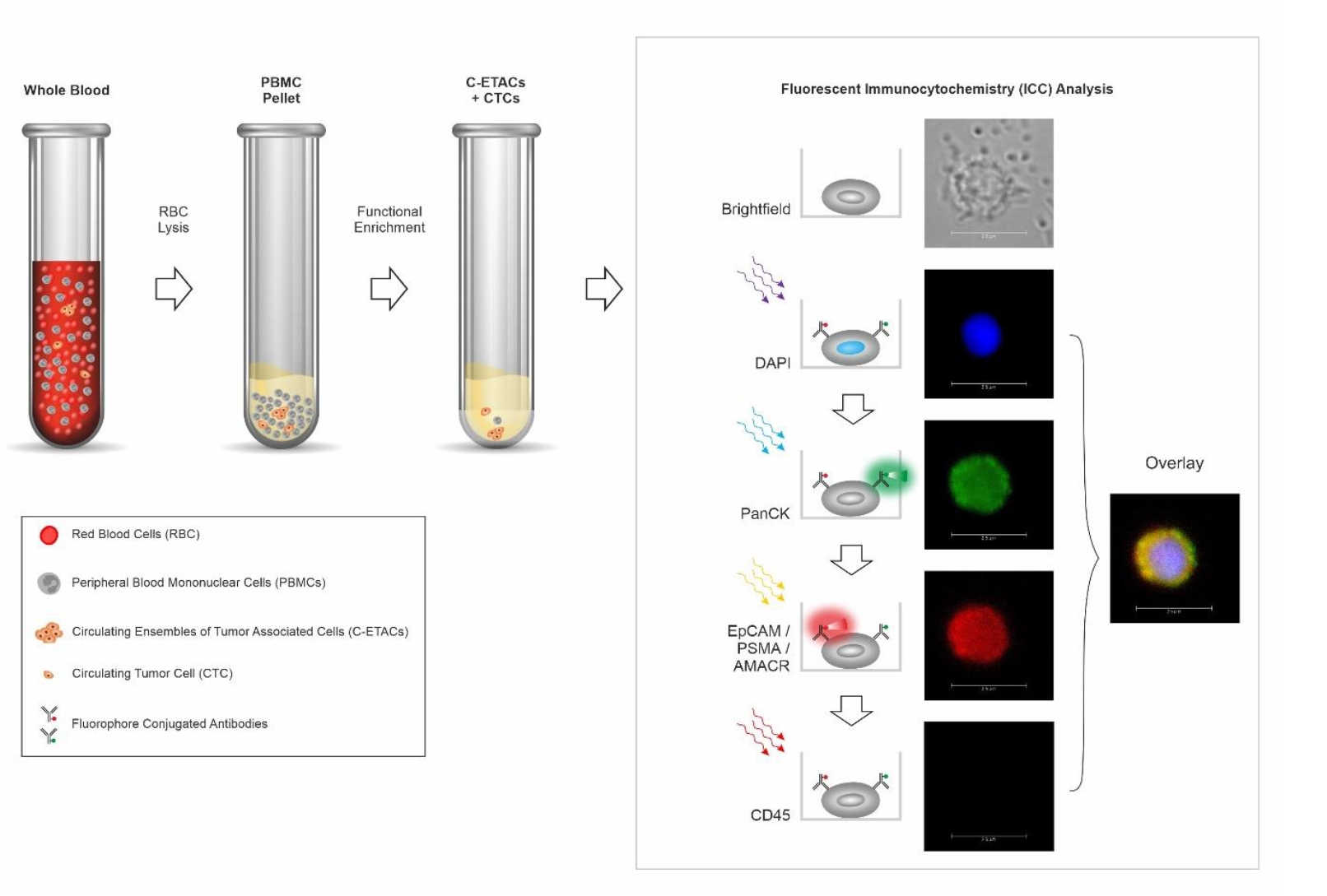
Schema of Test. Functional enrichment of CTCs is achieved using a proprietary CTC enrichment medium (CEM) that eliminates all non-malignant cells and permits tumor derived malignant cells to survive. Subsequently, the multiplexed immunocytochemistry (ICC) evaluates and identifies PrAD-CTCs based on positivity of the indicated markers.

**Figure 2.**
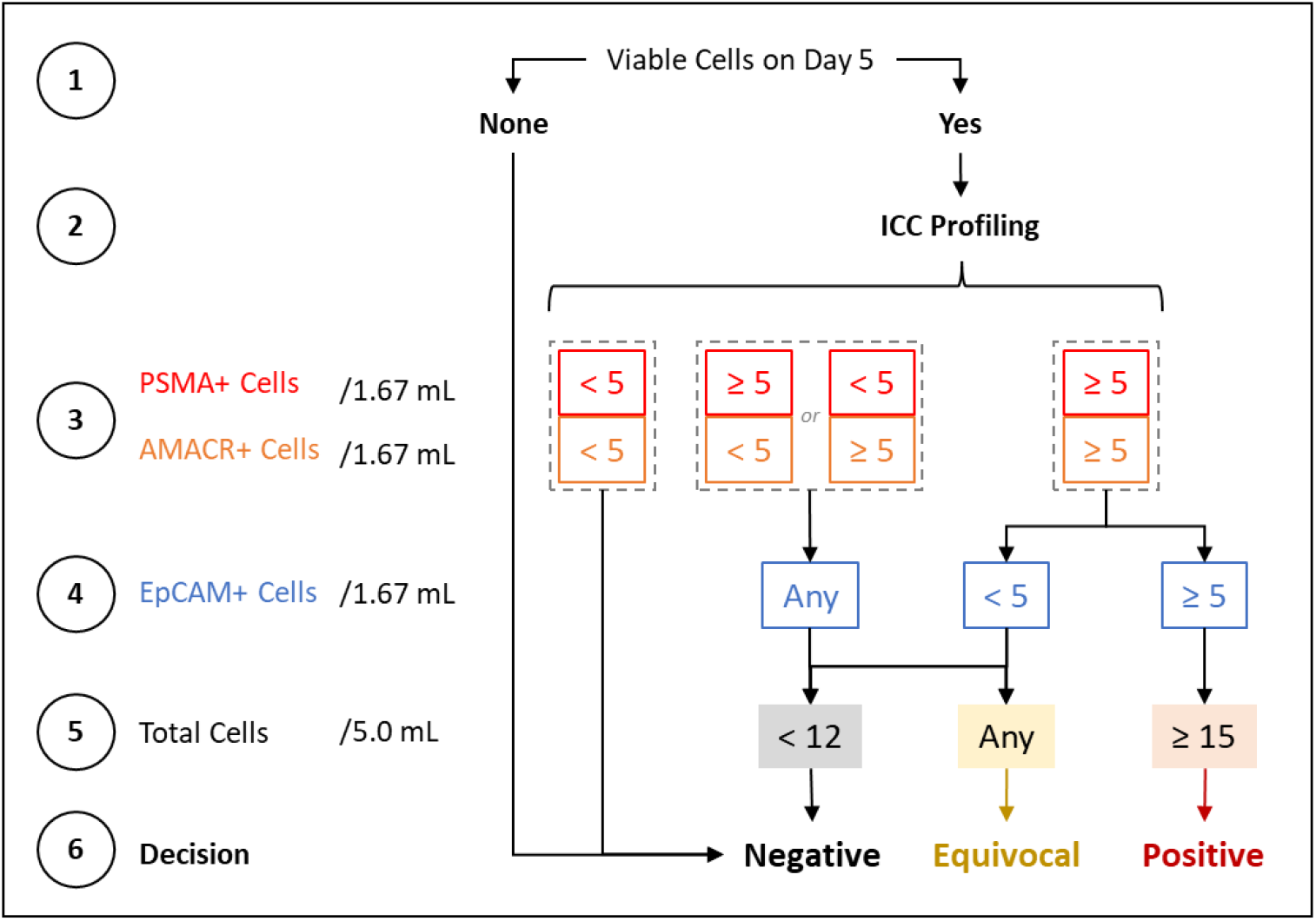
Decision Matrix for Classifying Samples. The detection threshold for PrAD-CTCs is ≥ 15 PanCK cells / 5 mL, which is constituted by the detection of ≥ 5 PSMA+ cells, ≥ 5 AMACR+ cells and ≥ 5 EpCAM+ cells in the respective aliquots. Priority is given to PSMA and AMACR over EpCAM while classifying samples as ‘**Positive**’ to ensure specificity for PrAD over other epithelial malignancies where EpCAM+ cells may be detected.

### Method Development and Optimization

Comprehensive details of method development and optimization studies as well as their findings are provided in the **Supplementary Materials**.

### Analytical Validation

Analytical validations were performed by determining the recovery of reference human prostate cancer cell line (VCaP) spiked into healthy donor blood samples. VCaP reference cells were spiked at various densities as per the design of and requirement for each validation parameter (specified in **Supplementary Materials**) into healthy donor blood samples, which were processed as per the test for enrichment of CTCs (spiked cells) and immunocytochemistry. The spike-recovery study design was applicable for validation of analyte stability (and recovery), linearity, limit of detection, limit of quantitation, limit of blank, sensitivity, specificity, accuracy, precision and interference. Comprehensive details of study design, observations and inferences are provided in **Supplementary Materials**.

### Case Control Clinical Study

The ability of the Test to discern / identify PrAD from asymptomatic males was first ascertained and established in a case control study with 160 males who were recently diagnosed, therapy naïve cases of PrAD and 800 healthy males aged 49 years and above with no prior diagnosis of any cancer, no current suspicion of any cancer and with serum PSA ≤ 0.5 ng / mL. Initially, samples in the Asymptomatic cohort were randomized into Training, Test and Validation Sets in a 60%:20%:20% ratio. The PrAD cases were first segregated by extent of disease as Localized (confined to primary site), Regional (spread to regional lymph nodes) and Distant (metastasized to distal lymph nodes or other organs) for which survival is known [2]. Subsequently, the samples were assigned to Training and Test Sets in a 60%:20%:20% ratio. The Training Set comprising of 96 PrAD and 480 healthy males’ samples was initially evaluated with the analysts unblinded to the status of the samples. Next the blinded Test Set comprising of 32 PrAD and 160 healthy males’ samples was evaluated prior to blinded evaluation of the 32 PrAD and 160 healthy males’ samples in the Validation Set. Subsequently all Training, Test and Validation set samples (PrAD and healthy) were shuffled and random 20% samples (extent-wise for PrAD) were selected for analysis as Validation Set Iteration 2. This shuffling step was repeated to obtain 20 iterations of the Validation Set from which median and range of Sensitivity, Specificity and Accuracy were reported.

### Prospective Clinical Study

The performance characteristics of the test was next ascertained and established in a prospective clinical study of blood samples from 210 males with urological symptoms who were suspected of PrAD based on either an enlarged prostate alone (n = 80) or an enlarged prostate in conjunction with clinically suspicious or significant (>3 ng / mL) serum PSA as determined by the Urologist (n = 130). All participants provided 5 mL blood sample prior to undergoing a prostate biopsy. The findings of the histopathological examination (HPE) and the final diagnosis (cancer or benign) were initially blinded to the sponsor. Clinical status of samples (cancer / benign) was revealed to sponsor only after sample analysis was complete and test findings shared with the clinical study investigator to determine Sensitivity, Specificity and Accuracy.

### Molecular Concordance Studies

In a combined subset of 20 samples from the case-control and prospective cohorts, where matched prostate cancer tumor tissue and blood samples were available, a molecular concordance study was performed. Tumor Tissue DNA (ttDNA) was isolated and profiled by Next Generation Sequencing (NGS) using the Ion Proton Platform and the Oncomine Comprehensive Assay v3 Panel to identify gene variants with loss of tumor suppression or gain of oncogenic function which have been previously reported to be significant in / associated with prostate cancer. Simultaneously, PBMCs were isolated from the matched blood samples and used for CTC enrichment. On the fifth day, genomic DNA (gDNA) isolated from all surviving cells was evaluated by a ddPCR assay specific to the detected gene variant on a BioRad QX200 platform. Concordance between tumor tissue and CTCs was determined as the proportion of the latter where the corresponding gene variant was detected by ddPCR.

Tissue samples from the same 20 patients were also evaluated by fluorescence in situ hybridization (FISH) as per manufacturer’s protocol for TMPRSS2-ERG fusion. In samples where tissue was positive for this variation, enriched and harvested CTCs were also evaluated by FISH for the same biomarker.

## RESULTS

### Method Development

The method development studies showed the viability of multiplexed fluorescence ICC for detection of PrAD CTCs even with significantly lower expression of EpCAM, PanCK, AMACR and PSMA than primary tumor cells or reference cells (**Supplementary Figure S1**), as well as other key aspects including specificity of marker combination to prostate cancer (**Supplementary Figure S2**), absence of PrAD CTCs in benign prostate conditions (**Supplementary Table S1**) and the ability of the test to detect CTCs irrespective of patient age (**Supplementary Figure S3**), PSA (**Supplementary Figure S4**) and Gleason Score (**Supplementary Figure S5**). Comprehensive details are provided in **Supplementary Materials**.

### Analytical Validation

**Table 1** is a summary of all the findings of the analytical validation study. Analytical validation established analyte stability (**Supplementary Table S2-S3**), demonstrated high sensitivity and specificity of the test (**Supplementary Table S4**), significant linear characteristics (**Supplementary Figure S6**), high precision (**Supplementary Table S5**) and no loss of sensitivity in presence of potentially interfering substances (**Supplementary Table S6**). Comprehensive details are provided in **Supplementary Materials**.

**Table 1.** Findings of Analytical Validation Studies. The summary of findings of the Analytical Validation Studies indicate that the Test provides consistent, accurate and reproducible results with little or no interference from routine endogenous or exogenous factors when samples are obtained, stored and processed under the recommended conditions.

|  | EpCAM,<br>PanCK, CD45 | PSMA,<br>PanCK, CD45 | AMACR,<br>PanCK, CD45 | Overall |
| --- | --- | --- | --- | --- |
| Analyte Stability | 48h |  |  |  |
| Recovery<sup>1</sup> | 97.2% | 94.4% | 94.4% | 91.7% |
| Limit of detection | < 1 cell / mL |  |  |  |
| Linear Range | 1-256 cells / mL |  |  |  |
| Linearity | R<sup>2</sup> ≥ 0.99 | R<sup>2</sup> ≥ 0.99 | R<sup>2</sup> ≥ 0.99 | R<sup>2</sup> ≥ 0.99 |
| Sensitivity | 95.0% | 92.5% | 92.5% | 92.5% |
| Specificity | 100.0% | 100.0% | 100.0% | 100.0% |
| Accuracy | 97.1% | 95.7% | 95.7% | 95.7% |
| Precision | CV ≤ 9% | CV ≤ 6% | CV ≤ 6% | CV ≤ 9% |
| Robustness | CV < 10% |  |  |  |
| <sup>1</sup>Above 10 cells / 5 mL as determined from the Linearity experiment. Values within parentheses represent 95% CI. |  |  |  |  |

### Clinical Studies

We evaluated the performance characteristics of the test in two clinical studies. The demographic details of the study cohorts are provided in **Supplementary Table S7**. Both studies were conducted in a South Asian cohort with <0.005% reported prostate cancer incidence [29], and also where the prostate cancer risk in asymptomatic males is significantly lower than the <7% reported among Caucasians with ≤ 0.5 ng / mL serum PSA [30,31] most of whom are also expected to be clinically insignificant prostate cancer [30,32]. Due to this low probability of an underlying prostate cancer in healthy subjects, they were a suitable ‘control’ population. Further, the selection of such a control population is also more ethical since it would be unethical to perform a biopsy on asymptomatic individuals for the sole purpose of ruling out prostate cancer for this study. The Case Control Study had a stringent, blinded, iterative cross-validation design which minimized the risk of overfitting. Detailed findings of the Training and Test Sets as well as the 20 iterations of the Validation sets are provided in **Supplementary Table S8**). In this study, the median sensitivity was 100% for local, regional and for metastatic disease as well as overall (**Table 2**). In absence of any positive or equivocal findings in the control (cancer free and asymptomatic) cohort, the specificity of the test (cancer v/s healthy) was 100%.

**Table 2.** Findings of Clinical Validation Studies. The table provides the Stage-wise and overall performance characteristics of the Test which were determined from 20 iterations of the Validation Set. Values within parentheses indicate 95% Confidence intervals.

|  | Case Control Study,<br>Cancer v/s Asymptomatic<br>*Specificity: 100.0% (95% CI: 97.7% - 100.0%)* |  | Prospective Study,<br>Cancer vs Benign<br>*Specificity: 100.0% (95% CI: 97.4% - 100.0%)* |  |
| --- | --- | --- | --- | --- |
|  | Sensitivity | Accuracy | Sensitivity | Accuracy |
| **Cumulative** | 100.0%<br>*95%CI: 89.1% - 100.0%* | 100.0%<br>*95%CI: 98.1% - 100.0%* | 91.2%<br>*95%CI: 81.8% - 96.7%* | 97.14%<br>*95%CI: 93.9% - 98.9%* |
| *Local* | 100.0%<br>*95%CI: 79.4% - 100.0%* | 100.0%<br>*95%CI: 97.9% - 100.0%* | 75.0%<br>*95%CI: 50.9% to 91.3%* | 96.9%<br>*95%CI: 92.9% - 98.9%* |
| *Regional* | 100.0%<br>*95%CI: 97.7% - 100.0%* | 100.0%<br>*95%CI: 97.8% - 100.0%* | 85.7%<br>*95%CI: 42.1% - 99.6%* | 99.3%<br>*95%CI: 96.3% - 99.9%* |
| *Distal* | 100.0%<br>*95%CI: 97.7% - 100.0%* | 100.0%<br>*95%CI: 97.8% - 100.0%* | 100.0%<br>*95%CI: 90.8% - 100.0%* | 100.0%<br>*95%CI: 97.9% - 100.0%* |

In the second (prospective) clinical study with 210 symptomatic males, 68 (32.4%) were eventually diagnosed with PrAD and 142 (67.6%) were diagnosed with various benign prostate conditions. There were no positive or equivocal findings among those diagnosed with benign prostate conditions. Hence the specificity of the test (cancer v/s benign) was 100%. Among the 68 cancer cases, the Test assigned 56 samples as positive, 6 as equivocal and 6 as negative (**Supplementary Table S9**), yielding a sensitivity of 91.2% since equivocals were considered as positive (**Table 2**). In the prospective study, the sensitivity of the test was observed to correlate positively with Gleason Scores and PSA levels (where available) (**Supplementary Table S10**).

### Molecular Concordance Studies

Among the 20 tumor samples tested, driver mutations with allele frequency were detected in 15 samples by NGS profiling of tumor tissue DNA using the Oncomine Comprehensive Assay v3 Panel on the Ion Proton Platform. Among these 15 patient samples, a specific TaqMan ddPCR assay was available for variants detected in 12 cases. CTC-enriched fraction from these samples were used for gDNA isolation which in turn was evaluated by a ddPCR assay specific to the driver mutation on a BioRad QX200 platform. Variants in ttDNA detected by NGS were also detected by ddPCR in 9 (75%) CTCs (**Supplementary Table S11**). A subset of 4 PrAD cases were identified where the tissue was positive for TMPRSS2-ERG fusion by FISH. The CTC enriched fraction from these 4 samples was evaluated by FISH and the TMPRSS2-ERG fusion was detected in 3 cases (75%). Overall, the orthogonal concordance studies appeared to confirm that the CTCs detected by the Test originated from the same prostate malignancy. The 75% concordance was considered satisfactory in light of clonal diversity in tumor cells and CTCs.

## DISCUSSION

We describe a blood test for Prostate cancer detection in asymptomatic males based on multiplexed fluorescence ICC profiling of CTCs functionally enriched from a 5 mL blood sample. The test can detect Prostate cancer irrespective of age, serum PSA level and Gleason grade and also has high sensitivity regardless of the extent of disease. Analytical validation ascertained accuracy and reliability of the test. The case control cross-validation study demonstrated 100% median overall sensitivity and 100.0% specificity for all stages of Prostate cancer. The subsequent prospective clinical validation study demonstrated 91.2% Sensitivity and 100% Specificity in the real world setting for detecting Prostate cancer and differentiating prostate cancer from benign prostate conditions. The Test has (a) high sensitivity for all stages, including early stages, (b) high specificity to minimize the risk of false positives, (c) high positive predictive value, (d) high negative predictive value. The performance characteristics of the test support its potential clinical utility in Prostate cancer screening among asymptomatic males.

Presently, evaluation of serum PSA is part of standard prostate cancer diagnostic work up in symptomatic men and is often evaluated as part of elective prostate cancer screening in asymptomatic males [33,34]. However, PSA testing has lower specificity and is associated with a high false positive rate, e.g. ∼66% [5]. Other PSA-based tests such as %-free PSA [35], [-2]pro-PSA (p2PSA) [36] and Prostate Health Index (PHI) [37] with documented sensitivity / specificity trade-off [35,38,39] are currently not recommended or approved for routine prostate cancer screening. The inverse relationship between specificity and sensitivity of PSA and PSA based tests [39] implies inefficient triaging where a significant proportion of individuals who do undergo a prostate biopsy based on these tests may actually be free from prostate cancer. Based on the limitations of serum PSA evaluations alone to provide meaningful insight into prostate cancer detection, Thompson et al suggested that ‘*PSA levels should no longer be referred to as ‘normal’ or ‘elevated’ but should be incorporated into a multivariable risk assessment to provide individualized risk information for decision making*’[40]. Among other non-invasive (blood-based) approaches, a pan-cancer detection test based on methylation profiling in cfDNA reported very low sensitivity (∼10%) for localized Prostate cancer [41,42].

Our test is based on detection of CTCs, which are ubiquitous in blood of patients with an underlying solid organ cancer [28] and unlikely in the blood of individuals without an underlying malignancy as well as those with other non-malignant or inflammatory conditions. CTCs are hence an ideal analyte to differentiate individuals with and without an underlying malignant condition with high specificity and sensitivity. There appear to be limited or no risks associated with use of the test since it is non- (or minimally) invasive and is performed on a venous draw of 5 mL peripheral blood. The potential benefits of the test include detection of Prostate cancer at early (localized) stages. The strengths of our study include (a) use of adequately powered sample sizes, (b) sample blinding to eliminate bias, (c) an iterative cross-validation design intended to eliminate risk of cross fitting, and (d) a prospective study in a real-world setting. The analytical and clinical validations described in this manuscript provide tangible evidence of the test performance which supports the hypothesis (design) as well as the intended use of the test. The high specificity translates into an exceedingly low risk of false positives in individuals with benign prostate conditions which eliminates or significantly reduces risks of overdiagnosis or overtreatment in these individuals.

Although the test has high performance characteristics for Prostate cancer detection, we note the following potential limitations of the test. Non-(adeno)-carcinoma types which account for <8% of Prostate cancer are not detected by this test. The sensitivity for the detection was lower for localized Prostate cancer in the prospective study (at ∼75%). We speculate that the false negative cases could be attributed to biologically different characteristics of some Prostate cancers. However, these false negatives would not add to the pre-existing risk of the individual since the risk of missing localized cancers can be partially mitigated by the higher sensitivity for subsequent detection at regional stages with comparable 5-year survival.

While the risk stratification of prostate cancer includes serum PSA level, clinical stage and Gleason score, a Gleason score of >8 is considered an independent predictor of high-risk disease with increased rates of treatment failures and poorer outcomes. Our test is not intended to provide information on, or correlate with, the Gleason score. Our test has high sensitivity for detection of high-grade / aggressive prostate cancers which require urgent multi-modality treatment approaches and where early detection is vital for more effective clinical management. There would appear to be a minimal risk of overdiagnosis from detection of low-grade (lower risk) prostate cancers which account for up to 66% of all prostate cancers [43]. However, since up to 40% of patients initially diagnosed with low-risk prostate cancer demonstrate pathological progression over time [44], detection of low-grade prostate cancers can benefit from active surveillance [45].

## CONCLUSION

The high sensitivity and specificity of the test enables prostate cancer detection and differentiation from benign prostate conditions (or healthy individuals) and presents significant advantages over PSA based approaches. The test has potential to reduce the need for invasive biopsies and thus significantly mitigates risks of overdiagnosis and overtreatment. The potential benefits of the test are compelling and support the need for further prospective large cohort clinical studies to determine the performance characteristics of the test for detection of prostate cancer, especially localized disease.

## Data Availability

All relevant data are included in the manuscript and its Supplementary Information file.

## ADDITIONAL INFORMATION

## ACKNOWLEDGEMENTS

The authors are grateful to the staff of the Study Sponsor (DCG) for their contributions in managing various clinical, operational and laboratory aspects of the study.

## ETHICS APPROVAL AND CONSENT TO PARTICIPATE

All biological samples were obtained from participants in three studies, TRUEBLOOD (http://ctri.nic.in/Clinicaltrials/pmaindet2.php?trialid=31879), ProState (http://ctri.nic.in/Clinicaltrials/pmaindet2.php?trialid=31713) and RESOLUTE (http://ctri.nic.in/Clinicaltrials/pmaindet2.php?trialid=30733). All studies were approved by Datar Cancer Genetics Limited Institutional Ethics Committee. All participants provided written informed consent. All studies were performed in accordance with the Declaration of Helsinki.

## DATA AVAILABILITY

All relevant data are included in the manuscript and its Supplementary Information file.

