## Supplementary file for "Accurate Prostate Cancer Detection based on Circulating Tumor Cell Profiling"

**SUPPLEMENTARY MATERIALS, METHODS AND FINDINGS**

**Markers**

The Test uses a multi-marker system which ensures the detection of Prostate Adenocarcinoma associated Circulating Tumor Cells (PrAD-CTCs) with high specificity, as malignant apoptosis resistant cells expressing (positive for) PanCK, PSMA, AMACR and EpCAM and negative for CD45. Pan-Cytokeratins (PanCK) are a family of cytoplasmic structural proteins expressed in epithelial tumors and CTCs. Common Leucocyte Antigen (CD45) as a negative marker serves to differentiate CTCs from CD45 positive haematolymphoid cells. Prostate Specific Membrane Antigen (PSMA) is a type II transmembrane glycoprotein, highly specific to and oncogenic in prostate adenocarcinomas. While PSMA is expressed in normal prostate tissue, it is overexpressed in prostate adenocarcinomas. Alpha Methylacyl CoA Racemase (AMACR) is a mitochondrial and peroxisomal enzyme, which is not expressed in normal prostate tissue but is overexpressed in prostate adenocarcinomas. Epithelial Cell Adhesion Molecule (EpCAM) are membrane antigens present on epithelial cells (and carcinomas) that function in cell adhesion. PSMA and AMACR are evaluated during routine diagnostic histopathological evaluation (HPE) of prostate tumor tissue. The PSMA+/AMACR+ combination ensures a high specificity of organ localisation.

**Antisera and Cell Lines**

The antisera used included recombinant human (RH) Anti CD326 IgG1-Vio 615 (Miltenyi Biotech), RH Anti-CK-IgG1-Vio 515 (Miltenyi Biotech), RH Anti-CD45-IgG1-APCVio 770 (Miltenyi Biotech), Mouse Anti-PSMA IgG1 (Dako), Rabbit Anti-AMACR IgG1 (Dako), Anti-Mouse Alexa Fluor 594 (Invitrogen) and Anti-Rabbit Alexa Fluor 594 (Invitrogen). The reference cell lines include VCaP (human prostate adenocarcinoma), MOLT-3 (leukemia) and SW982 (synovial sarcoma) all of which were procured from ATCC. The purity of reference cell lines was confirmed by Short Tandem Repeat (STR) Profiling and testing for Mycoplasma every 6 months.

**Isolation of Primary Tumor Derived Cells**

Tissue samples with >70% tumor content (as confirmed by HPE) were dissociated into single-cell suspensions using a combination of mechanical dissociation and enzymatic degradation of the extracellular matrix using the Tumor Cell Isolation Kit, human kit components and the gentleMACS™ Dissociator (Miltenyi Biotech, Germany). The single cell suspension of Tumor Derived Cells (TDCs) obtained by this method was cultured at 37°C under 5% CO_2_ and 4% O_2_ for 24 hours after which viable cells (TDCs) were harvested by gentle centrifugation (400 × *g*, 5 min, 4°C) and resuspended in PBS.

**Enrichment of Circulating Tumor Cells from Peripheral Blood**

Aliquoted blood samples (5 mL) were processed for the enrichment of circulating tumor cells (CTCs) from peripheral blood mononuclear cells (PBMC) as described previously [1].Briefly, PBMCs were isolated from whole blood via lysis of red blood cells (RBCs) followed by centrifugation. PBMCs were resuspended in Phosphate Buffered Saline (PBS) and treated with a proprietary CTC enrichment medium (CEM) that induces cell death in all apoptosis-competent non-malignant (hemato-lymphoid, epithelial and endothelial) cells, while malignant tumor derived cells (CTCs) survive due to apoptosis resistance. After treatment for 5 days at 37°C, surviving cells and cell clusters are harvested by gentle centrifugation (400 × *g*, 5 min, 4°C) and resuspended in PBS.

**Immunocytochemistry Profiling of CTCs**

Briefly, CTCs enriched from 5 mL of blood were resuspended in 1500 μL 1x Phosphate Buffered Saline (PBS) and 100 μL aliquots of enriched CTCs seeded into 15 wells. Cells in each well were equivalent to 333 μL blood sample. Cells were fixed with 4 % Paraformaldehyde, Permeabilized with 0.3% Triton-X 100 and treated with 3% BSA (blocking). Cells were immunostained with each of the 3 separate Primary (1°) Ab cocktails for multiplexed analysis of the following combination of markers, (a) Anti-PanCK (1:500), Anti-CD45 (1:500), Anti-EpCAM (1:500), (b) Anti-PanCK (1:500), Anti-CD45 (1:500), Anti-PSMA (1:2), (c) Anti-PanCK (1:500), Anti-CD45 (1:500), Anti-AMACR (1:4). Samples for PSMA and AMACR are incubated with secondary (2°) anti-mouse Ab (1:500). PBS washes followed each Ab incubation step. Each marker combination was evaluated in 5 wells (333 μL × 5 = 1.67 mL equivalent of blood). Finally, cells were treated with 4’,6-Diamidino-2-phenylindole dihydrochloride (DAPI) for nuclear staining. Control samples (VCaP for EpCAM, PSMA and AMACR; MOLT3 for CD45) were included in each run. Samples were evaluated on the CellInsight High Content Screening (HCS) Platform to determine the Fluorescence Intensity (FI) for each marker. Marker expression was determined by the sequential excitation and acquisition of fluorescence signal.

Cells isolated from a primary benign or malignant tumor were resuspended in phosphate buffered saline (PBS) solution and ICC profiled similarly as described above for CTCs.

**Method Development**

*Detection Thresholds*

Study: VCaP cells, MOLT-3 cells, SW982 cells, PrAD-CTCs, PrAD-TDCs (M-TDCs) and TDCs from known benign prostate tumor (B-TDCs) were immunostained for all markers. FI was recorded for each marker in respective channels and recorded to determine differences in expression level per cell type.

Findings: The FI of PSMA, AMACR, EpCAM and PanCK were significantly higher in VCaP, M-TDC and PrAD-CTCs than MOLT-3, SW982 and B-TDC (**Supplementary Figures S1**). Conversely, the FI of CD45 was higher in MOLT-3 than the other cell types. The FI threshold for positivity was assigned as 60,000 Relative Fluorescence Units (RFU) for PanCK and EpCAM and 50,000 RFU for PSMA and AMACR. These thresholds accommodate CTCs with lower marker expression, e.g., CTCs undergoing EMT. For CD45 40,000 RFU was set as the lower threshold for MOLT-3 (PC for CD45) and as the upper threshold for VCaP (NC for CD45) and PrAD-CTCs. In addition to FI, numerical thresholds were defined for acceptance of PC and NC as the minimum proportion of cells staining positively in PC (>60%) and NC (<1%) for each marker.

*Marker Specificity*

Study: We determined the specificity of the marker combination to PrC by evaluating the expression (FI) of PSMA and AMACR were evaluated in CTCs from Cancers of the Lung, Kidney, Bone / Sarcoma, Colon, Head and Neck and Pancreas.

Findings: Expression of PSMA and AMACR were lower (FI < 50,000 U) in all non-PrC CTCs (**Supplementary Figure S2**).

*Marker Expression in Prostate Cancer*

Study: FI for PSMA, AMACR, EpCAM, PanCK and CD45 were evaluated in subsets of PrAD-CTCs stratified by Age (n = 73), Serum PSA level at diagnosis (n = 90) and Gleason Grade Group (n = 72).

Findings: There was no suppression in FI of any marker based on age-group, serum PSA or Gleason Grade Group indicating that the test can detect PrAD-CTCs irrespective of these prognostic variables (**Supplementary Figures S3 – S5**).

*PrAD-CTCs in Non-malignant Prostate Conditions*

Study: To determine the Specificity of the Test to discern PrC from non-malignant conditions of the Prostate, we evaluated blood samples from 260 individuals who were recently diagnosed with Benign Prostate Hyperplasia or Prostatitis. Samples were processed for CTC enrichment and ICC profiling as described above.

Findings: Among the blood samples from 260 known cases of (**Supplementary Table S1**) benign prostate conditions, PanCK+, EpCAM+, CD45- cells were detected in 5 samples, of which CTC in 1 sample were also positive for PSMA but not AMACR. Based on the absence of PrAD-CTC (PanCK+, EpCAM+, PSMA+, AMACR+, CD45-) in all samples, the test had a Specificity of 100%.

**Analytical Validation**

Analytical validations were performed by determining the recovery of VCaP cells spiked into healthy donor blood samples. Table 1 is a summary of all the findings of the analytical validation study. Analytical validation established analyte stability, demonstrated high sensitivity and specificity of the test, significant linear characteristics, high precision and no loss of sensitivity in presence of potentially interfering substances. Findings of analytical validation which established these performance characteristics of the test are provided in the Supplementary Materials.

*Stability and Recovery*

Study: To establish the analyte stability, 27 × 5 mL aliquots of healthy donor blood were spiked with ~15 VCaP cells each (final = 3 cells / mL) and were stored at 2°C - 8°C. Of the 27 aliquots, 9 aliquots were processed immediately and 9 aliquots each were processed after 24 h and after 48 h respectively. Of the 9 aliquots evaluated at each time point, 3 aliquots each were used to determine recovery of PSMA+, AMACR+ and EpCAM+ cell types respectively. In addition, 3 × 5 mL aliquots of blood were collected from 5 known PrAD cases; one aliquot from each patient was processed immediately (0 h), while the other 2 aliquots were stored at 2°C - 8°C and processed after 24 h and 48 h respectively. Recoveries at 0 h were normalized as 100% and recoveries at 24 h and 48 h were determined relative to the 0 h recovery.

Findings: In the spiked samples, >93.3% recovery was observed for each cell type for up to 48 h (**Supplementary Table S2**). In clinical samples, the overall (combined PanCK+) recovery was 95.3% and 82.8% at 24h and 48h respectively, when 0 h recovery was normalized as 100% (**Supplementary Table S3**). The findings of the stability and recovery study indicated that clinical samples could be stored at 2°C-8°C for up to 48h with <20% loss of cells.

*Linearity*

Study: VCaP cells were spiked into 264 × 5 mL aliquots of healthy donor blood, stored for 48 h at 2°C - 8°C and then processed. The 264 aliquots comprised 3 sets of 88 aliquots (11 spike densities × 8 replicates) – each set was assigned to either of the 3 multiplexed marker combinations. The study also included 24 × 5 mL aliquots (3 sets × 8 replicates) of healthy donor blood samples which were not spiked. Samples were stored for 48 h at 2°C - 8°C. Linearity was evaluated by Linear Regression.

Findings: Recoveries of spiked cells were generally higher above 5 cells / 5 mL (**Supplementary Figure S6**). R^2^ ≥0.99 in all markers demonstrated the linear response characteristics of the method, especially in the range of 5 - 1280 cells / 5 mL (1 – 256 cells / mL) which is considered the reportable range.

*Limits of Detection, Quantitation and Blank*

Study: The Limit of Blank (LoB) was determined from the 24 × 5 mL unspiked healthy donor blood samples in the Linearity study. The Limit of Detection (LoD) was determined from a subset of the Linearity Study which included 72 × 5 mL samples spiked with 1, 3 or 5 VCAP cells (24 of each). The Limit of Quantitation (LoQ) was determined from a subset of the Linearity Study which included 96 × 5 mL samples spiked with 1, 3, 5 or 10 VCAP cells (24 of each).

Findings:

No PSMA+, AMACR+ or EpCAM+ cells were detected in any of the unspiked samples (no false positives). Thus, the limit of blank was determined to be 0 cells / mL. The Limit of Detection (LoD) was determined as 1 cell / 5 mL. The limit of quantitation (LoQ) was 10 cells / 5 mL (2 cells / mL).

*Sensitivity, Specificity and Accuracy*

Study: VCaP cells were spiked into 40 × 15 mL aliquots (5 spikes × 8 replicates) of healthy donor blood at 15, 30, 60, 120 and 240 cells. Each 15 mL sample was split into 3 × 5 mL aliquots that were used for the analysis of each of the 3 marker combinations. Samples were stored for 48 h at 2°C - 8°C prior to analysis. Unspiked healthy donor blood samples (24 × 5 mL) were included in this study for the determination of Specificity. Samples with equivocal findings were considered as negative. Accuracy was determined based on total true positive and true negative samples detected out of the total 70 samples.

Findings: Among the 40 spiked samples evaluated for sensitivity, VCaP cells were detected in 37 samples, yielding a sensitivity of 92.5%. Since VCaP cells were undetectable in any of the 30 un-spiked samples, the specificity was deemed to be 100%. Accuracy was determined to be 95.7%. (**Supplementary Table S4**).

*Precision*

Study: On Day 1, User 1 spiked 15 (Low) VCaP cells into each of 8 × 5 mL aliquots of healthy donor blood, and 150 (High) VCaP cells into each of another 8 × 5 mL aliquots of healthy donor blood. All samples (8 Low spike + 8 High) were stored for 48 h at 2°C - 8°C and processed. User 1 performed this study on 10 consecutive days and used one of two HCS Instruments. User 2 independently replicated the study on 10 consecutive days and used the second HCS Instrument. Mean Recoveries (%) were used to calculate Standard Deviation (SD) and Coefficient of Variation (CV, %).

Intra-Run, Inter-Run and Inter-Operator Precision were determined.

Findings: **Supplementary Table S5** provides the intra-run, inter-run and inter-operator %CV for low spike, high spike and the cumulative, respectively. The cumulative %CV was ≤6.1% for intra-run, ≤1.9% for inter-operator and ≤2.4% for inter-run. The overall %CV was ≤8.3% indicating high precision of the test.

*Interfering Substances*

Study: The performance characteristics of the Test were evaluated in presence of endogenous (serum markers) and exogenous (non-anticancer drugs) factors as possible interfering agents (**Supplementary Table S6**). The drugs selected for evaluation of interference represent the most commonly prescribed non-anticancer medications in the US and Europe. The endogenous factors also represent commonly observed variables during blood pathology work-up. Analytical grade molecules were used to prepare working stock solutions and immediately used for spiking studies. All drugs were evaluated at the reported Peak Plasma Concentrations (C_Max_), while serum markers were evaluated at concentrations that are considered as elevated. Blood from a healthy donor (75 mL) who was not under any medication (last 14 days) was procured from a blood bank and spiked with about 750 VCaP cells. The spiked sample was split into 25 × 3 mL aliquots; 21 aliquots were spiked with each of the above substances at the indicated concentrations and 4 aliquots were used as unspiked controls. Each 3 mL sample was split into 3 × 1 mL aliquots; one aliquot each was used for detection of PanCK+ EpCAM+ cells, PanCK+ PSMA+ cells and PanCK+ AMACR+ cells respectively.

Findings: The presence of non-anticancer drugs at medically relevant peak plasma concentrations (C_Max_) or the serum parameters evaluated did not impact the recovery or detection of VCaP cells spiked into blood samples. The study established the ability of the test to remain unaffected in presence of systemic treatment agents (drugs) and elevated serum parameters.

**SUPPLEMENTARY FIGURES**

**Supplementary Figure S1. Detection Thresholds.** The expression levels (FI: fluorescence intensity) of PSMA (A), AMACR (B), EpCAM (C), PanCK (D) and CD45 (E) were evaluated on VCaP (reference human prostate adenocarcinoma cell line) MOLT-3 (leukemia cell line), SW982 (sarcoma cell line), benign prostate tumor derived cells (B-TDC), prostate AD (malignant) tumor derived cells (M-TDC) and prostate AD CTCs (PrAD-CTC). The expression levels were used to assign detection thresholds for positivity for each marker,

**Supplementary Figure S2. PSMA and AMACR Expression in various non-Prostate cancer CTCs.** (A) PSMA in various CTCs, (B) AMACR in various CTCs.

**Supplementary Figure S3. Age-group and Marker Expression on CTCs.** (A) PSMA, (B) AMACR, (C) EpCAM, (D) PanCK, (E) CD45.

**Supplementary Figure S4. Preoperative Serum PSA level and Marker Expression on CTCs.** (A) PSMA, (B) AMACR, (C) EpCAM, (D) PanCK, (E) CD45.

**Supplementary Figure S5. Gleason Grade Group and Marker Expression on CTCs.** (A) PSMA, (B) AMACR, (C) EpCAM, (D) PanCK, (E) CD45.

**Supplementary Figure S6: Analytical Validation: Linearity.** The Test exhibited significant linearity with R^2^ ≥0.99. The Tabulated values below the figure show the recovery and range of recovery.

**SUPPLEMENTARY TABLES**

**Supplementary Table S1. Non-malignant Prostate Conditions evaluated for Specificity.** Stratification of samples.

**Supplementary Table S2. Analytical Validation Stability and Recovery of Spiked Cells.** VCaP cells were spiked into healthy donor blood samples and the recovery of spiked cells was evaluated for up to 48 hours.

**Supplementary Table S3. Analytical Validation Stability and Recovery of CTCs in Clinical Samples.** Blood samples from known PrAD-CTC positive cases were evaluated for recovery of PrAD CTCs for up to 48 hours.

**Supplementary Table S4. Analytical Validation: Sensitivity, Specificity, Accuracy.** VCaP cells were spiked into healthy donor blood samples at various seed densities and their recoveries evaluated to determine Sensitivity. Unspiked healthy donor blood samples were evaluated for false positives to determine Specificity. Accuracy was determined from Sensitivity and Specificity.

**Supplementary Table S5. Analytical Validation: Precision.** Recovery of low spiked VCaP cells in healthy donor blood samples across multiple replicates by 2 independent operators and over multiple days were used to determine the %CV.

**Supplementary Table S6. Analytical Validation: Impact of Potentially Interfering Substances.** The Test was not prone to interference from endogenous agents (deranged serum parameters) and exogenous agents (common non-anticancer drugs)

**Supplementary Table S7. Demographics of Clinical Study Participants**

The table provides demographic details such as age, serum PSA and Gleason Grade Group (prostate cancer cases) for all study participants whose samples were used for the validation study.

**Supplementary Table S8. Case Control Cross Validation Study Findings.**

The table shows the number and % of samples in Training, Test and the 20 iterations of the Validation Sets with Positive, Equivocal and Negative findings.

**Supplementary Table S9. Prospective Study Findings.**

The table shows the number and % of samples with Positive, Equivocal and Negative findings.

**Supplementary Table S10. CTC detection based on PSA and Gleason Score.**

The table shows the Prospective Study samples with Positive, Equivocal and Negative findings against a PSA v/s Gleason Score Stratification Matrix

**Supplementary Table S11. Molecular Concordance Study Findings.**

The table shows the number of samples where ddPCR on CTC-DNA were positive for the significant gene variant identified by NGS profiling of tumor tissue DNA.

**Supplementary Figure S1. Detection Thresholds.**

(A) PSMA

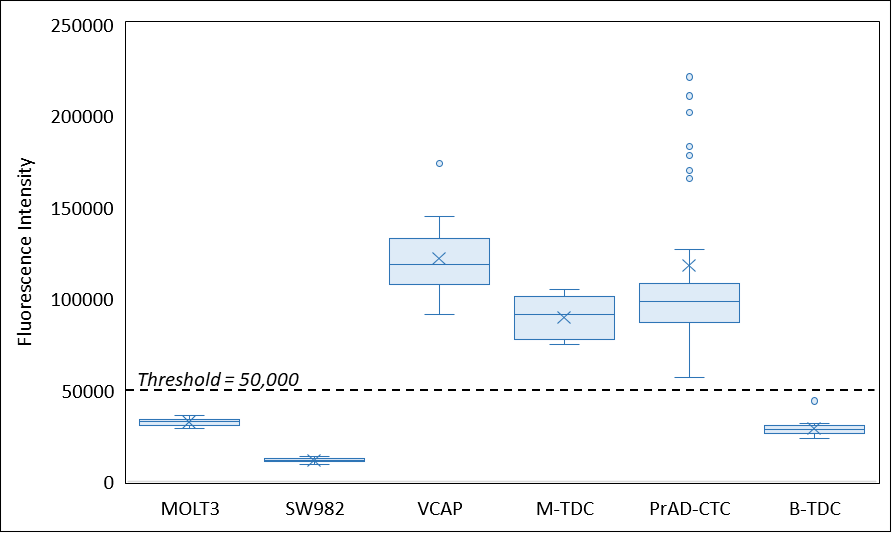

(B) AMACR

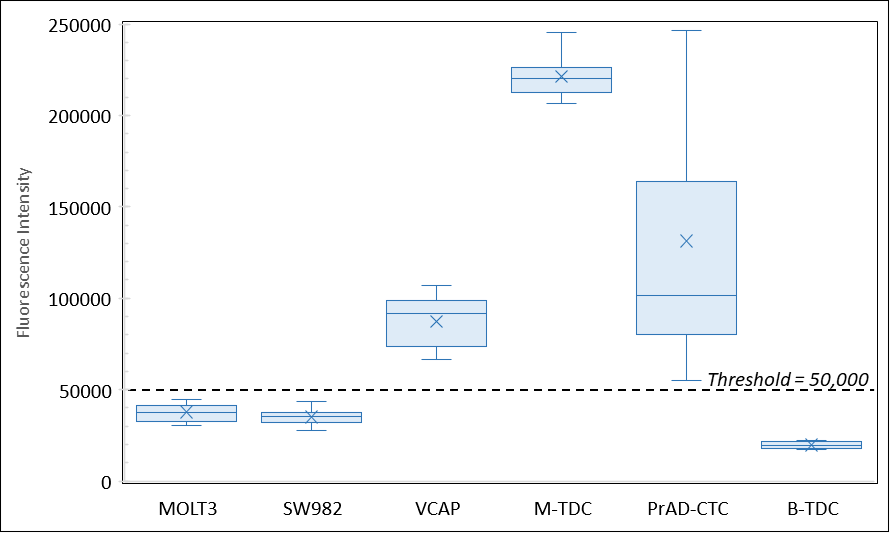

(C) EpCAM

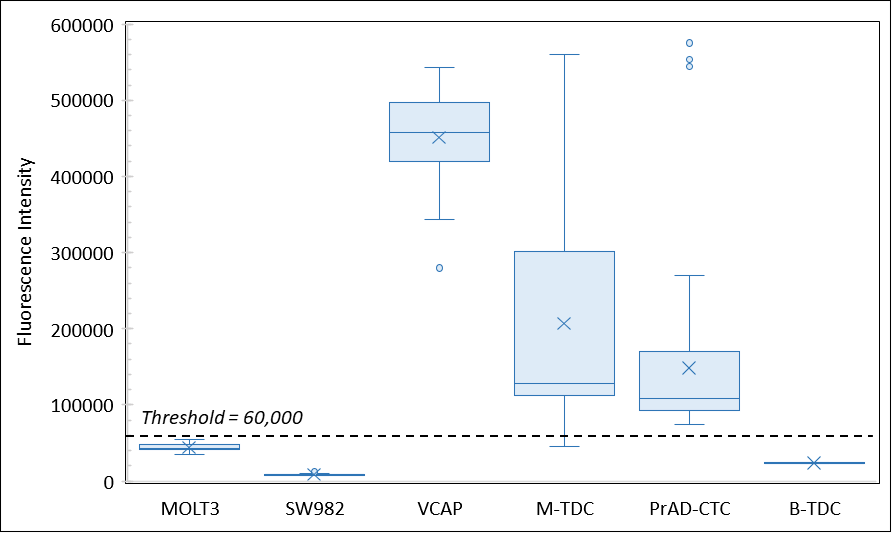

(D) PanCK

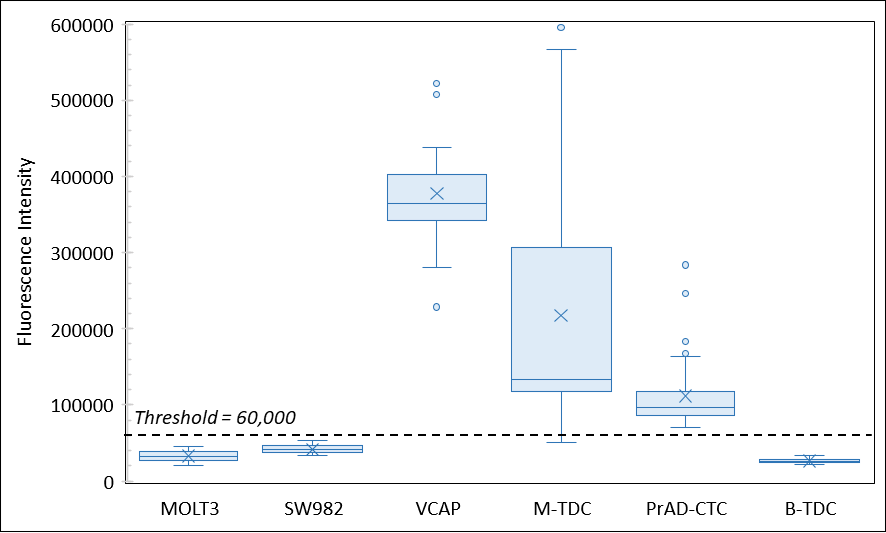

(E) CD45.

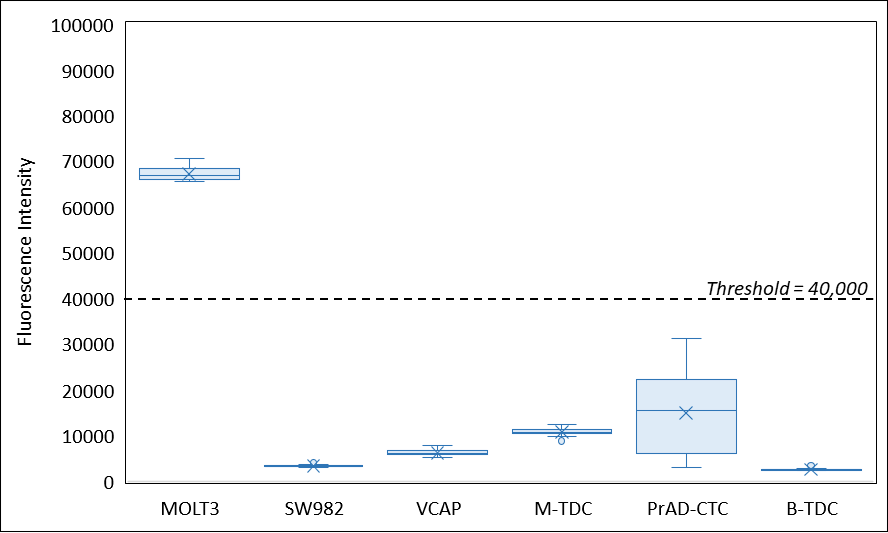

**Supplementary Figure S2. PSMA (A) and AMACR (B) Expression in various CTCs.**

(A)

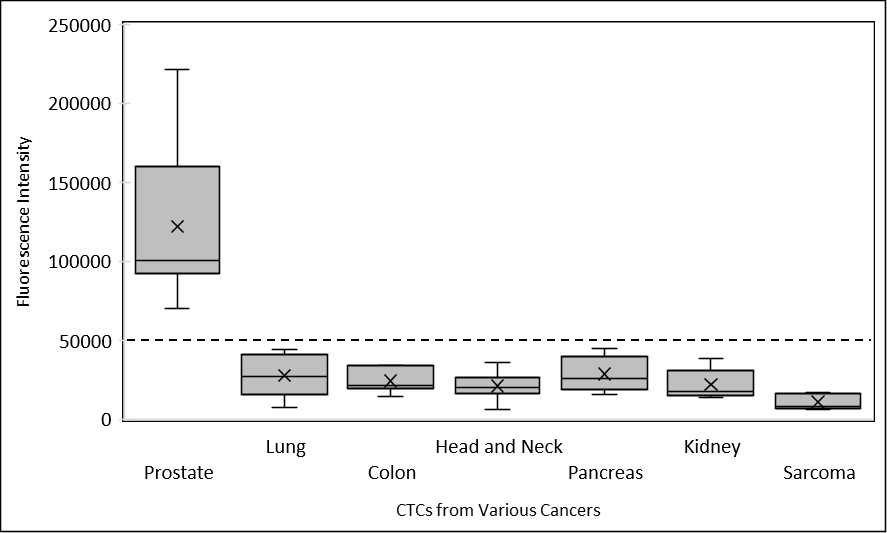

(B)

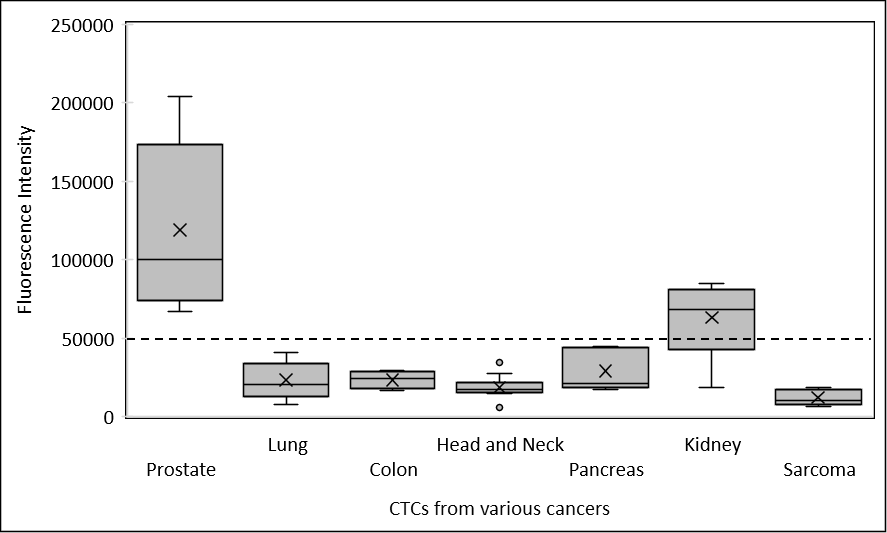

**Supplementary Figure S3. Age-group and Marker Expression on CTCs.**

(A) PSMA

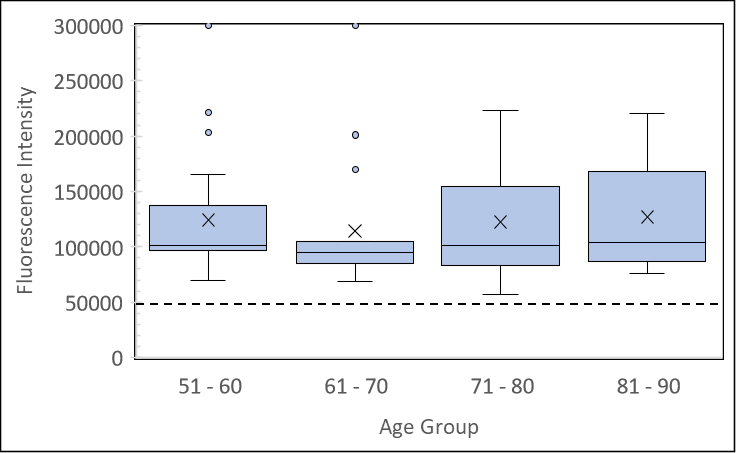

(B) AMACR

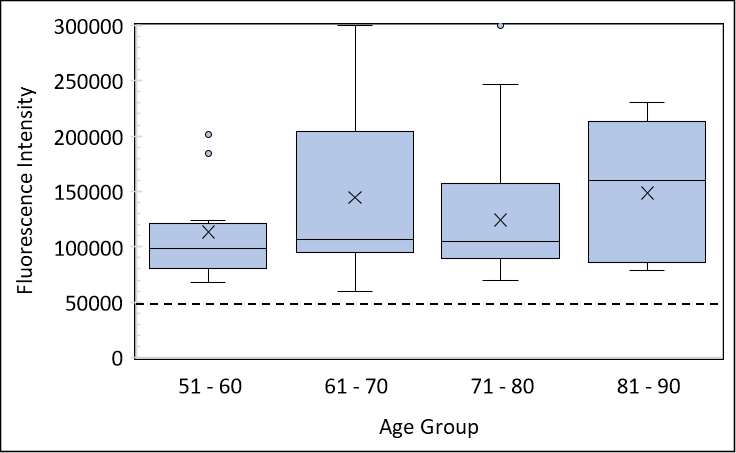

(C) EpCAM

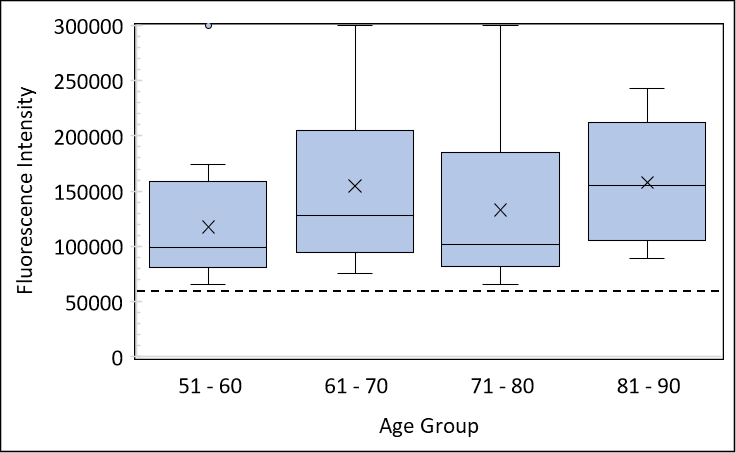

(D) PanCK

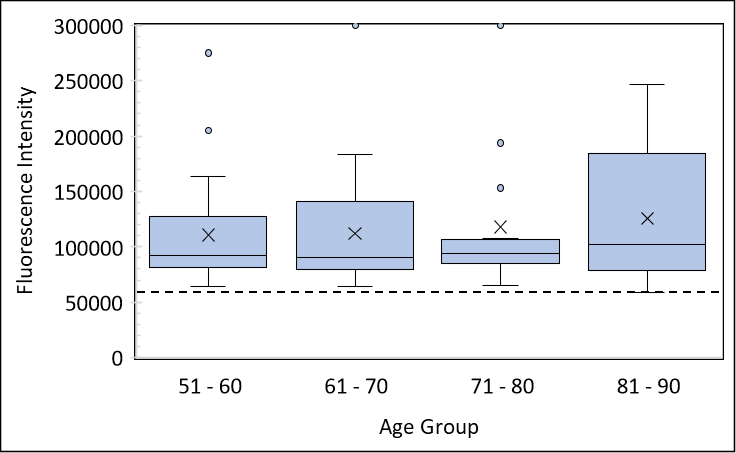

(E) CD45.

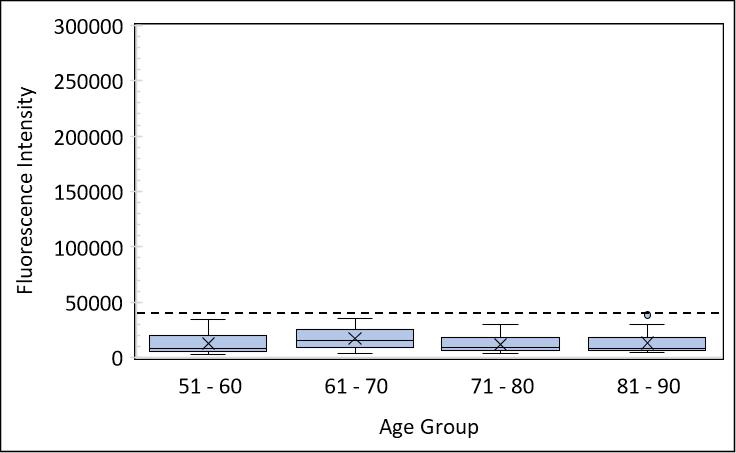

**Supplementary Figure S4. Serum PSA level and Marker Expression on CTCs.**

A. PSMA

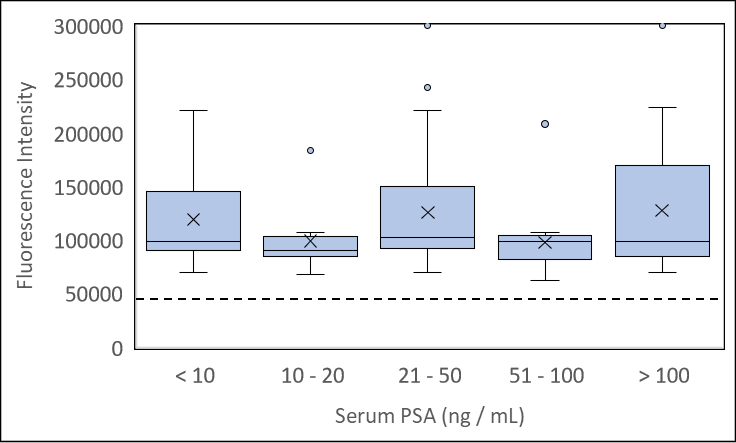

B. AMACR

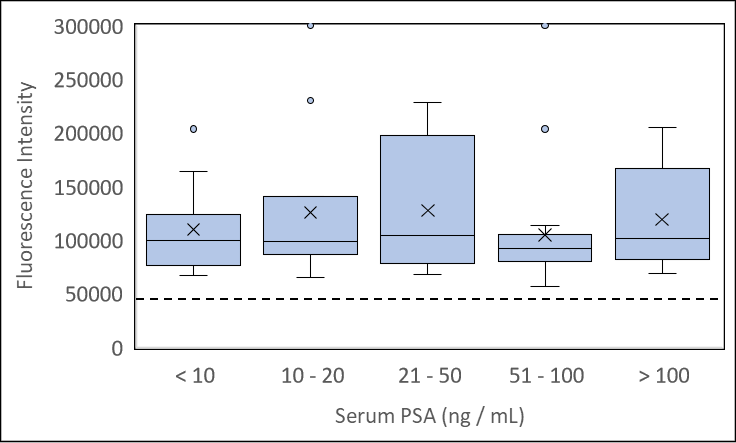

C. EpCAM

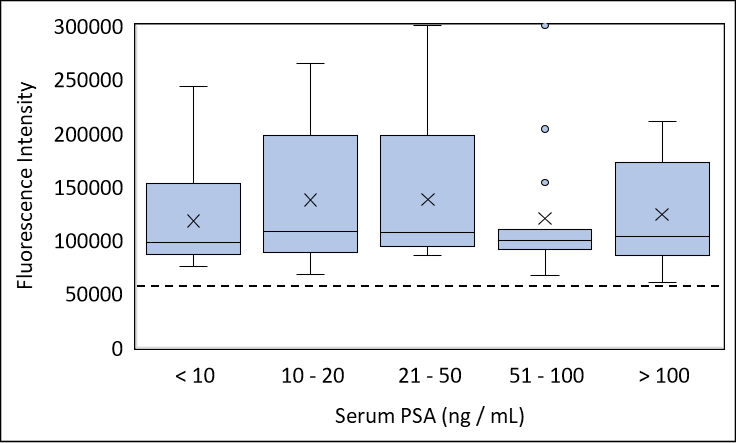

D. PanCK

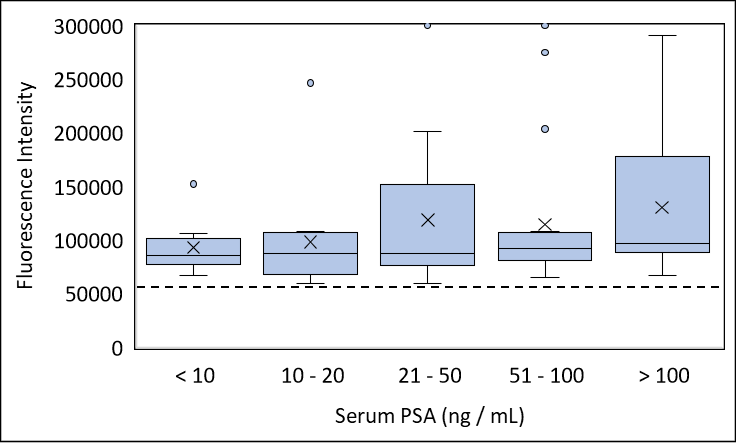

E. CD45

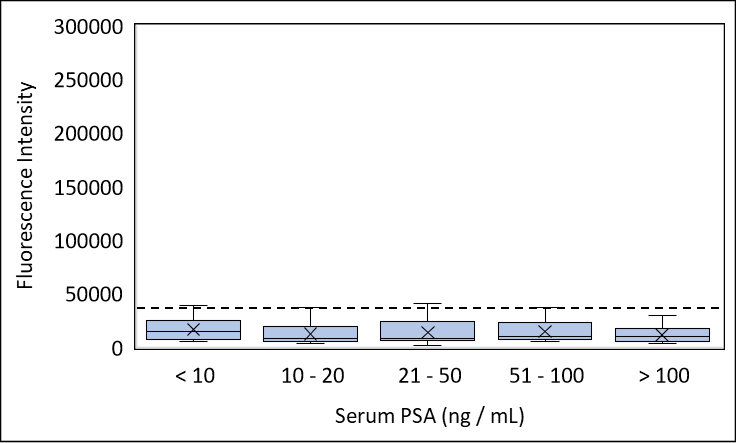

**Supplementary Figure S5. Gleason Grade Group and Marker Expression on CTCs.**

A. PSMA

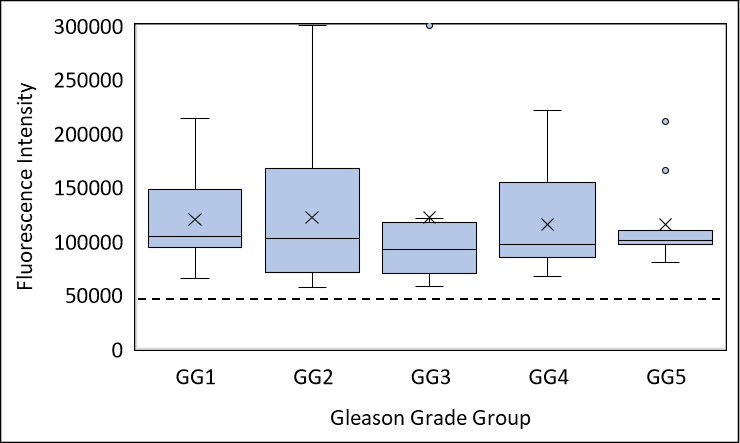

B. AMACR

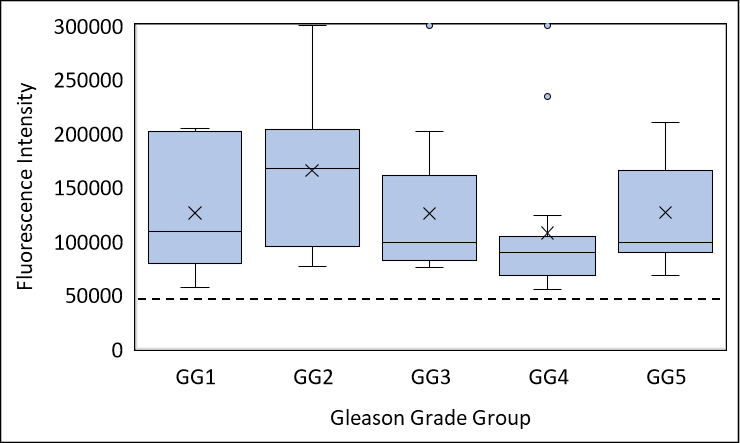

C. EpCAM

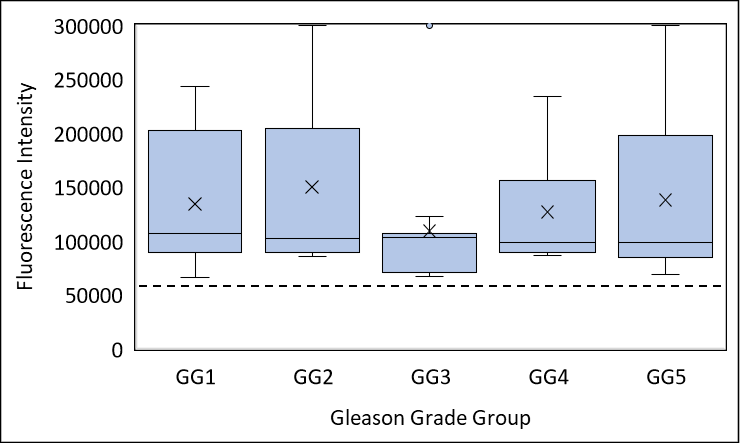

D. PanCK

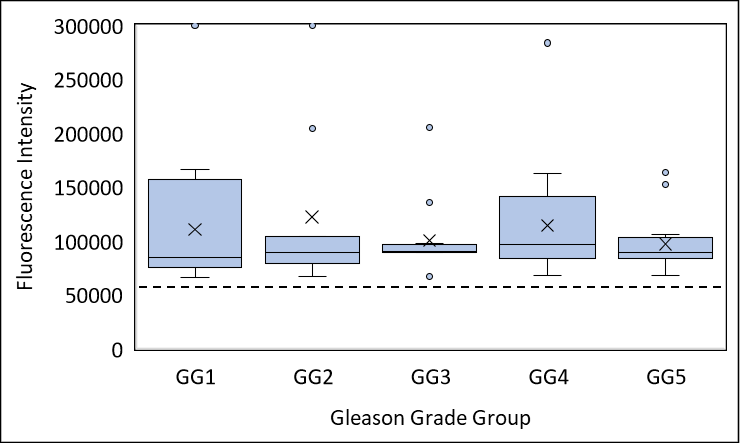

E. CD45

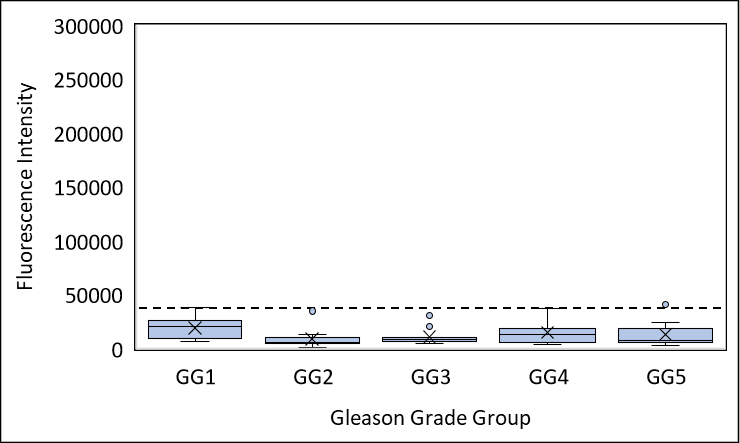

**Supplementary Figure S6: Analytical Validation: Linearity.**

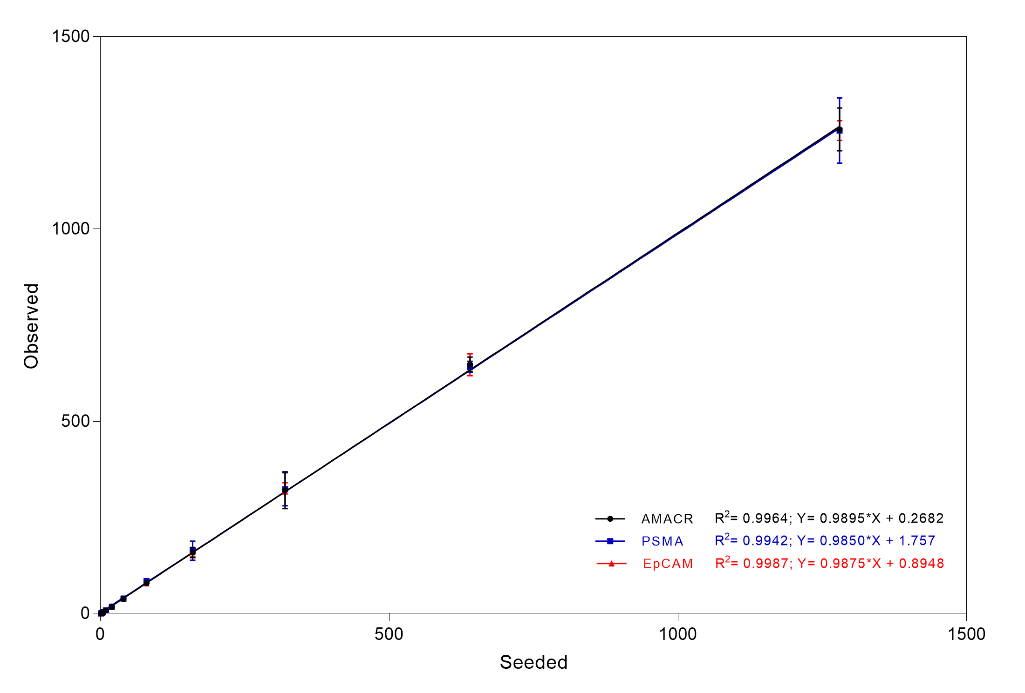

| **Spiked cells** | **Mean and Range of Detected Cell Counts (8 Replicates)** | | |
| --- | --- | --- | --- |
|  | **PanCK+,**  **EpCAM+** | **PanCK+,**  **PSMA+** | **PanCK+,**  **AMACR+** |
| 1250 | 1256  (1213 – 1284) | 1256  (1132 – 1372) | 1259  (1138 – 1310) |
| 640 | 647  (595 – 678) | 641  (626 – 669) | 648  (615 – 680) |
| 320 | 326  (305 – 344) | 323  (273 – 385) | 321  (259 – 376) |
| 160 | 159  (146 – 178) | 164  (141 – 198) | 159  (138 – 174) |
| 80 | 79  (72 - 86) | 84  (78 - 88) | 79  (73 - 87) |
| 40 | 38  (35 - 39) | 39  (34 - 45) | 39  (31 - 45) |
| 20 | 19  (16 – 22) | 18  (12 – 22) | 17  (13 – 25) |
| 10 | 9  (7 - 10) | 8  (7 - 9) | 8  (6 - 11) |
| 5 | 4  (2 – 5) | 4  (2 – 6) | 4  (3 – 6) |
| 3 | 2  (0 – 3) | 2  (1 – 4) | 2  (1 – 3) |
| 1 | 0  (0 – 1) | 0  (0 – 1) | 0  (0 – 1) |
| 0 | 0  0 – 0 | 0  0 – 0 | 0  0 – 0 |

**SUPPLEMENTARY TABLES**

**Supplementary Table S1. Non-malignant Prostate Conditions.**

| **Parameter** | **Value** |
| --- | --- |
| **Median Age (Range)** | 67 years (38 – 90 years) |
| **Diagnosis**  Hyperplasia  Hypertrophy  Prostatitis  Reactive Atypia  Squamous Metaplasia | 119  2  137  1  1 |

**Supplementary Table S2. Analytical Validation Stability and Recovery of Spiked Cells.**

| **Time (h)** | **Spiked Cells** | **Mean Recovery, % Recovery and Recovery Range (%)** | | |
| --- | --- | --- | --- | --- |
|  |  | **PanCK+, EpCAM+** | **PanCK+, AMACR+** | **PanCK+, PSMA+** |
| 0 | 15 | 16 (106.7%)  (100.0% - 113.3%) | 15 (100%)  (100.0% - 106.7%) | 15 (100%)  (100.0% - 106.7%) |
| 24 | 15 | 15 (100.0%)  (100.0% - 100.0%) | 14 (93.3%)  (93.3% - 100.0%) | 14 (93.3%)  (93.3% - 100.0%) |
| 48 | 15 | 14 (93.3%)  (93.3% - 100.0%) | 14 (93.3%)  (93.3% - 100.0%) | 14 (93.3%)  (93.3% - 93.3%) |
| 72 | 15 | 13 (86.7%)  (80.0% - 93.3%) | 13 (86.7%)  (80.0% - 93.3%) | 13 (86.7%)  (80.0% - 93.3%) |
| 96 | 15 | 12 (80.0%)  (73.3% - 86.7%) | 11 (73.3%)  (66.7% - 80.0%) | 12 (80.0%)  (73.3% - 86.7%) |
| 120 | 15 | 11 (73.3%)  (66.7% - 80.0%) | 11 (73.3%)  (66.7% - 80.0%) | 11 (73.3%)  (66.7% - 80.0%) |

**Supplementary Table S3. Analytical Validation Stability and Recovery of CTCs in Clinical Samples.**

| **Time (h)** | **Cell Types, Detected Numbers, % Recovery and % Recovery Range** | | | | | | | | |
| --- | --- | --- | --- | --- | --- | --- | --- | --- | --- |
|  | **Patient** | **PanCK+, EpCAM+**  **cells (a)** | | **PanCK+, AMACR+**  **cells (b)** | | **PanCK+,**  **PSMA+**  **cells (c)** | | **Total PanCK+ cells**  **(a + b + c)** | |
| **0 h** | P_1_ | 6 | ***100%*** | 6 | ***100%*** | 5 | ***100%*** | 17 | ***100%*** |
|  | P_2_ | 5 |  | 5 |  | 5 |  | 15 |  |
|  | P_3_ | 8 |  | 6 |  | 6 |  | 20 |  |
|  | P_4_ | 5 |  | 9 |  | 5 |  | 19 |  |
|  | P_5_ | 7 |  | 5 |  | 5 |  | 17 |  |
| **24 h** | P_1_ | 5 | 83.3% | 5 | 83.3% | 5 | 100.0% | 15 | 88.2% |
|  | P_2_ | 4 | 80.0% | 5 | 100.0% | 5 | 100.0% | 14 | 93.3% |
|  | P_3_ | 7 | 87.5% | 6 | 100.0% | 6 | 100.0% | 19 | 95.0% |
|  | P_4_ | 5 | 100.0% | 9 | 100.0% | 5 | 100.0% | 19 | 100.0% |
|  | P_5_ | 7 | 100.0% | 5 | 100.0% | 5 | 100.0% | 17 | 100.0% |
|  | Mean  (Range) | 90.2%  (80.0% - 100%) | | 96.7%  (83.3% - 100%) | | 100.0%  (100% - 100%) | | 95.3%  (88.2% - 100%) | |
| **48 h** | P_1_ | 5 | 83.3% | 5 | 83.3% | 4 | 80.0% | 14 | 82.4% |
|  | P_2_ | 4 | 80.0% | 4 | 80.0% | 4 | 80.0% | 12 | 80.0% |
|  | P_3_ | 7 | 87.5% | 5 | 83.3% | 5 | 83.3% | 17 | 85.0% |
|  | P_4_ | 4 | 80.0% | 8 | 88.9% | 4 | 80.0% | 16 | 84.2% |
|  | P_5_ | 6 | 85.7% | 4 | 80.0% | 4 | 80.0% | 14 | 82.4% |
|  | Mean  (Range) | 83.3%  (80.0% - 87.5%) | | 83.1%  (80.0% - 88.9%) | | 80.7%  (80.0% - 83.3%) | | 82.8%  (80.0% - 85.0%) | |

**Supplementary Table S4. Analytical Validation: Sensitivity, Specificity, Accuracy.**

| **Sample Type** | **Number of cells** | | | **Samples** | | |
| --- | --- | --- | --- | --- | --- | --- |
|  | **Spiked** | **Detected:**  **Mean, Range** | **Total** | | **Negative** | **Positive** |
| **PanCK+, AMACR+** | | | | | | |
| **Unspiked** | - | - | **30** | | **30 (100%)** | - |
| **Spiked** | **(overall)**  **5**  **10**  **20**  **40**  **80** | -  4 (3-6)  8 (6-11)  17 (13-25)  39 (31 – 45)  79 (73 – 87) | **40**  8  8  8  8  8 | | **2**  2  -  -  -  - | **38 (95.0%)**  6  8  8  8  8 |
| **PanCK+, PSMA+** | | | | | | |
| **Unspiked** | - | - | **30** | | **30 (100%)** | - |
| **Spiked** | **(overall)**  **5**  **10**  **20**  **40**  **80** | -  4 (2 – 6)  8 (7 – 9)  18 (12 - 22)  39 (34 – 45)  84 (78 – 88) | **40**  8  8  8  8  8 | | **3**  3  -  -  -  - | **37 (92.5%)**  5  8  8  8  8 |
| **PanCK+, EpCAM+** | | | | | | |
| **Unspiked** | - | - | **30** | | **30 (100%)** | - |
| **Spiked** | **(overall)**  **5**  **10**  **20**  **40**  **80** | -  4 (2 – 5)  9 (7 – 10)  19 (16 – 22)  38 (35 – 39)  79 (72 – 86) | **40**  8  8  8  8  8 | | **3**  3  -  -  -  - | **37 (92.5%)**  5  8  8  8  8 |
| **Overall PanCK+** | | | | | | |
| **Unspiked** | - | - | **30** | | **30 (100%)** | - |
| **Spiked** | **(overall)**  **5**  **10**  **20**  **40**  **80** | -  12 (8 – 16)  25 (21 – 28)  54 (50 – 65)  116 (107 – 123)  242 (231 – 254) | **40**  8  8  8  8  8 | | **3**  3  -  -  -  - | **37 (92.5%)**  5  8  8  8  8 |

**Supplementary Table S5. Analytical Validation: Precision.**

|  | **Low Spike (15 cells / 5 mL)** | | | **High Spike (150 cells / 5 mL)** | | | **Overall CV%** |
| --- | --- | --- | --- | --- | --- | --- | --- |
|  | **Mean** | **SD** | **CV%** | **Mean** | **SD** | **CV%** |  |
| **PanCK+, AMACR+, CD45- cells** | | | | | | | |
| ***Intra-Run*** | | | | | | | |
| Operator 1 | *15.2* | *1.16* | ***7.7%*** | *153.5* | *6.78* | ***4.4%*** | ***6.0%*** |
| Operator 2 | *15.4* | *1.15* | ***7.5%*** | *153.4* | *7.00* | ***4.6%*** | ***6.0%*** |
| Cumulative | *15.3* | *1.07* | ***7.0%*** | *153.5* | *6.54* | ***4.3%*** | ***5.6%*** |
| ***Inter-Run*** | | | | | | | |
| Operator 1 | *15.2* | *0.46* | ***3.0%*** | *153.5* | *1.61* | ***1.1%*** | ***2.0%*** |
| Operator 2 | *15.4* | *0.38* | ***2.5%*** | *153.4* | *1.88* | ***1.2%*** | ***1.8%*** |
| Cumulative | *15.3* | *0.22* | ***1.4%*** | *153.5* | *1.45* | ***0.9%*** | ***1.2%*** |
| ***Inter-Operator*** | | | | | | | |
| Inter-Operator | *15.3* | *0.16* | ***1.0%*** | *153.5* | *0.04* | ***0.0%*** | ***0.5%*** |
| **OVERALL** | *-* | *-* | ***7.6%*** | - | - | ***4.5%*** | ***6.0%*** |
| **PanCK+, PSMA+, CD45- cells** | | | | | | | |
| ***Intra-Run*** | | | | | | | |
| Operator 1 | *15.3* | *1.13* | ***7.4%*** | *153.1* | *7.17* | ***4.7%*** | ***6.0%*** |
| Operator 2 | *15.2* | *1.13* | ***7.4%*** | *151.9* | *5.99* | ***3.9%*** | ***5.7%*** |
| Cumulative | *15.3* | *1.07* | ***7.0%*** | *152.5* | *6.08* | ***4.0%*** | ***5.5%*** |
| ***Inter-Run*** | | | | | | | |
| Operator 1 | *15.3* | *0.35* | ***2.3%*** | *153.1* | *2.56* | ***1.7%*** | ***2.0%*** |
| Operator 2 | *15.2* | *0.25* | ***1.7%*** | *151.9* | *1.76* | ***1.2%*** | ***1.4%*** |
| Cumulative | *15.3* | *0.18* | ***1.2%*** | *152.5* | *1.70* | ***1.1%*** | ***1.2%*** |
| ***Inter-Operator*** | | | | | | | |
| Inter-Operator | *15.3* | *0.04* | ***0.3%*** | *152.5* | *0.82* | ***0.5%*** | ***0.4%*** |
| **OVERALL** | *-* | *-* | ***7.4%*** | *-* | *-* | ***4.3%*** | ***5.9%*** |
| **PanCK+, EpCAM+, CD45- cells** | | | | | | | |
| ***Intra-Run*** | | | | | | | |
| Operator 1 | *16.6* | *1.62* | ***9.8%*** | *154.7* | *8.60* | ***5.6%*** | ***7.7%*** |
| Operator 2 | *16.0* | *1.88* | ***11.7%*** | *156.5* | *8.91* | ***5.7%*** | ***8.7%*** |
| Cumulative | *16.3* | *1.15* | ***7.0%*** | *155.6* | *8.05* | ***5.2%*** | ***6.1%*** |
| ***Inter-Run*** | | | | | | | |
| Operator 1 | *16.6* | *1.11* | ***6.6%*** | *154.7* | *3.48* | ***2.2%*** | ***4.4%*** |
| Operator 2 | *16.0* | *1.57* | ***9.8%*** | *156.5* | *2.98* | ***1.9%*** | ***5.9%*** |
| Cumulative | *16.3* | *0.58* | ***3.6%*** | *155.6* | *1.87* | ***1.2%*** | ***2.4%*** |
| ***Inter-Operator*** | | | | | | | |
| Inter-Operator | *16.3* | *0.47* | ***2.9%*** | *155.6* | *1.26* | ***0.8%*** | ***1.9%*** |
| **OVERALL** | - | - | ***10.9%*** | - | - | ***5.6%*** | ***8.3%*** |

**Supplementary Table S6. Analytical Validation: Impact of Potentially Interfering Substances.**

| **Agent** | **Concentration** | **Detected Cells / mL** | | |
| --- | --- | --- | --- | --- |
|  |  | **PanCK+,**  **EpCAM+** | **PanCK+,**  **AMACR+** | **PanCK+,**  **PSMA** |
| Lisinopril | 58 ng / mL | 8 (80%) | 9 (90%) | 8 (80%) |
| Atorvastatin | 30 ng / mL | 8 (80%) | 10 (100%) | 8 (80%) |
| Metformin | 5 μg / mL | 9 (90%) | 9 (90%) | 9 (90%) |
| Amlodipine | 5 ng / mL | 10 (100%) | 8 (80%) | 8 (80%) |
| Metoprolol | 50 ng / mL | 9 (90%) | 10 (100%) | 10 (100%) |
| Omeprazole | 660 ng / mL | 9 (90%) | 8 (80%) | 9 (90%) |
| Albuterol | 4.2 ng / mL | 10 (100%) | 9 (90%) | 10 (100%) |
| Ranitidine | 450 ng / mL | 8 (80%) | 9 (90%) | 9 (90%) |
| Azithromycin | 500 ng / mL | 9 (90%) | 8 (80%) | 8 (80%) |
| Paracetamol | 9.9 μg / mL | 8 (80%) | 10 (100%) | 10 (100%) |
| Aspirin | 3 μg / mL | 10 (100%) | 8 (80%) | 9 (90%) |
| Loperamide | 3.4 ng / mL | 9 (90%) | 9 (90%) | 9 (90%) |
| Dextromethorphan | 2.9 ng / mL | 8 (80%) | 9 (90%) | 8 (80%) |
| Sildenafil Citrate | 440 ng / mL | 8 (80%) | 8 (80%) | 9 (90%) |
| Cortisone | 2700 nM | 8 (80%) | 8 (80%) | 10 (100%) |
| Cholesterol | 3 mg / mL | 9 (90%) | 8 (80%) | 9 (90%) |
| Uric Acid | 150 μg / mL | 9 (90%) | 9 (90%) | 10 (100%) |
| Bilirubin | 20 μg / mL | 10 (100%) | 9 (90%) | 8 (80%) |
| Haemoglobin | 200 mg / mL | 10 (100%) | 8 (80%) | 9 (90%) |
| Glucose | 3 mg / mL | 9 (90%) | 10 (100%) | 8 (80%) |
| EDTA | 20 mg / mL | 8 (80%) | 9 (90%) | 9 (90%) |
| Control | - | 10 (100%) | 10 (100%) | 10 (100%) |

**Supplementary Table S7. Demographics of Clinical Study Participants**

| **Study Type** | **Case Control Study** | | **Prospective Study** | |
| --- | --- | --- | --- | --- |
| **Sample Type** | **Cancer** | **Healthy** | **Cancer** | **Benign** |
| **N =** | **160** | **800** | **68** | **142** |
| **Age (years)**  Median  Range | 70  (47 – 92) | 56  (48 – 75) | 68  (50 – 84) | 66  (25 – 90) |
| **Cancer Stage**  Local (T_1-3_N_0_M_0_)  Regional (T_x_N_1_M_0_)  Distal (T_x_N_x_M_1_)  n.a. | 80  40  40  - | - | 20  7  38  3 | - |
| **PSA (ng / mL)**  ≤ 0.5  0.5 – 3.0  3.0 – 10.0  10.0 – 20.0  > 20  n.a. | 3  7  14  13  97  26 | 800  -  -  -  -  - | 1  2  7  8  36  14 | 5  59  49  12  17  - |
| **Gleason Group**  1  2  3  4  5  n.a. | 17  26  16  28  29  44 | - | 8  6  6  17  8  23 | - |
| *n.a.: not available* | | | | |

**Supplementary Table S8. Case Control Cross Validation Study Findings.**

| ***Iteration*** | ***Sample*** | ***Samples*** | ***Negative*** | | ***Equivocal*** | | ***Positive*** | |
| --- | --- | --- | --- | --- | --- | --- | --- | --- |
|  | **Training Set** | | | | | | | |
| - | **Asymptomatic** | **480** | **480** | **100.0%** |  |  |  | **0.0%** |
|  | **Cancer** | **96** | **1** | 1.0% | **4** | 4.2% | **91** | 94.8% |
|  | *Local* | 48 | 1 | 2.1% | 2 | 4.2% | 45 | 93.8% |
|  | *Regional* | 24 |  | 0.0% | 2 | 8.3% | 22 | 91.7% |
|  | *Distal* | 24 |  | 0.0% |  | 0.0% | 24 | 100.0% |
|  | **Test Set** | | | | | | | |
| - | **Asymptomatic** | **160** | **160** | **100%** |  |  |  |  |
|  | **Cancer** | **32** | **1** | 3.1% | **0** | 0.0% | **31** | 96.9% |
|  | *Local* | 16 | 1 | 6.3% |  | 0.0% | 15 | 93.8% |
|  | *Regional* | 8 |  | 0.0% |  | 0.0% | 8 | 100.0% |
|  | *Distal* | 8 |  | 0.0% |  | 0.0% | 8 | 100.0% |
|  | **Validation Set** | | | | | | | |
| 1 | **Asymptomatic** | **160** | **160** | **100%** |  |  |  |  |
|  | **Cancer** | **32** | **0** | 0.0% | **1** | 3.1% | **31** | 96.9% |
|  | *Local* | 16 |  | 0.0% |  | 0.0% | 16 | 100.0% |
|  | *Regional* | 8 |  | 0.0% | 1 | 12.5% | 7 | 87.5% |
|  | *Distal* | 8 |  | 0.0% |  | 0.0% | 8 | 100.0% |
| 2 | **Asymptomatic** | **160** | **160** | **100%** |  |  |  |  |
|  | **Cancer** | **32** | **0** | 0.0% | **1** | 3.1% | **31** | 96.9% |
|  | *Local* | 16 |  | 0.0% | 1 | 6.3% | 15 | 93.8% |
|  | *Regional* | 8 |  | 0.0% |  | 0.0% | 8 | 100.0% |
|  | *Distal* | 8 |  | 0.0% |  | 0.0% | 8 | 100.0% |
| 3 | **Asymptomatic** | **160** | **160** | **100%** |  |  |  |  |
|  | **Cancer** | **32** | **1** | 3.1% | **0** | 0.0% | **31** | 96.9% |
|  | *Local* | 16 | 1 | 6.3% |  | 0.0% | 15 | 93.8% |
|  | *Regional* | 8 |  | 0.0% |  | 0.0% | 8 | 100.0% |
|  | *Distal* | 8 |  | 0.0% |  | 0.0% | 8 | 100.0% |
| 4 | **Asymptomatic** | **160** | **160** | **100%** |  |  |  |  |
|  | **Cancer** | **32** | **2** | 6.3% | **1** | 3.1% | **29** | 90.6% |
|  | *Local* | 16 | 2 | 12.5% |  | 0.0% | 14 | 87.5% |
|  | *Regional* | 8 |  | 0.0% | 1 | 12.5% | 7 | 87.5% |
|  | *Distal* | 8 |  | 0.0% |  | 0.0% | 8 | 100.0% |
| 5 | **Asymptomatic** | **160** | **160** | **100%** |  |  |  |  |
|  | **Cancer** | **32** | **0** | 0.0% | **0** | 0.0% | **32** | 100.0% |
|  | *Local* | 16 |  | 0.0% |  | 0.0% | 16 | 100.0% |
|  | *Regional* | 8 |  | 0.0% |  | 0.0% | 8 | 100.0% |
|  | *Distal* | 8 |  | 0.0% |  | 0.0% | 8 | 100.0% |
| 6 | **Asymptomatic** | **160** | **160** | **100%** |  |  |  |  |
|  | **Cancer** | **32** | **1** | 3.1% | **0** | 0.0% | **31** | 96.9% |
|  | *Local* | 16 |  | 0.0% |  | 0.0% | 16 | 100.0% |
|  | *Regional* | 8 | 1 | 12.5% |  | 0.0% | 7 | 87.5% |
|  | *Distal* | 8 |  | 0.0% |  | 0.0% | 8 | 100.0% |
| 7 | **Asymptomatic** | **160** | **160** | **100%** |  |  |  |  |
|  | **Cancer** | **32** | **1** | 3.1% | **1** | 3.1% | **30** | 93.8% |
|  | *Local* | 16 | 1 | 6.3% |  | 0.0% | 15 | 93.8% |
|  | *Regional* | 8 |  | 0.0% | 1 | 12.5% | 7 | 87.5% |
|  | *Distal* | 8 |  | 0.0% |  | 0.0% | 8 | 100.0% |
| 8 | **Asymptomatic** | **160** | **160** | **100%** |  |  |  |  |
|  | **Cancer** | **32** | **0** | 0.0% | **1** | 3.1% | **31** | 96.9% |
|  | *Local* | 16 |  | 0.0% |  | 0.0% | 16 | 100.0% |
|  | *Regional* | 8 |  | 0.0% | 1 | 12.5% | 7 | 87.5% |
|  | *Distal* | 8 |  | 0.0% |  | 0.0% | 8 | 100.0% |
| 9 | **Asymptomatic** | **160** | **160** | **100%** |  |  |  |  |
|  | **Cancer** | **32** | **1** | 3.1% | **0** | 0.0% | **31** | 96.9% |
|  | *Local* | 16 | 1 | 6.3% |  | 0.0% | 15 | 93.8% |
|  | *Regional* | 8 |  | 0.0% |  | 0.0% | 8 | 100.0% |
|  | *Distal* | 8 |  | 0.0% |  | 0.0% | 8 | 100.0% |
| 10 | **Asymptomatic** | **160** | **160** | **100%** |  |  |  |  |
|  | **Cancer** | **32** | **0** | 0.0% | **0** | 0.0% | **32** | 100.0% |
|  | *Local* | 16 |  | 0.0% |  | 0.0% | 16 | 100.0% |
|  | *Regional* | 8 |  | 0.0% |  | 0.0% | 8 | 100.0% |
|  | *Distal* | 8 |  | 0.0% |  | 0.0% | 8 | 100.0% |
| 11 | **Asymptomatic** | **160** | **160** | **100%** |  |  |  |  |
|  | **Cancer** | **32** | **0** | 0.0% | **1** | 3.1% | **31** | 96.9% |
|  | *Local* | 16 |  | 0.0% |  | 0.0% | 16 | 100.0% |
|  | *Regional* | 8 |  | 0.0% | 1 | 12.5% | 7 | 87.5% |
|  | *Distal* | 8 |  | 0.0% |  | 0.0% | 8 | 100.0% |
| 12 | **Asymptomatic** | **160** | **160** | **100%** |  |  |  |  |
|  | **Cancer** | **32** | **0** | 0.0% | **1** | 3.1% | **31** | 96.9% |
|  | *Local* | 16 |  | 0.0% |  | 0.0% | 16 | 100.0% |
|  | *Regional* | 8 |  | 0.0% | 1 | 12.5% | 7 | 87.5% |
|  | *Distal* | 8 |  | 0.0% |  | 0.0% | 8 | 100.0% |
| 13 | **Asymptomatic** | **160** | **160** | **100%** |  |  |  |  |
|  | **Cancer** | **32** | **0** | 0.0% | **0** | 0.0% | **32** | 100.0% |
|  | *Local* | 16 |  | 0.0% |  | 0.0% | 16 | 100.0% |
|  | *Regional* | 8 |  | 0.0% |  | 0.0% | 8 | 100.0% |
|  | *Distal* | 8 |  | 0.0% |  | 0.0% | 8 | 100.0% |
| 14 | **Asymptomatic** | **160** | **160** | **100%** |  |  |  |  |
|  | **Cancer** | **32** | **0** | 0.0% | **0** | 0.0% | **32** | 100.0% |
|  | *Local* | 16 |  | 0.0% |  | 0.0% | 16 | 100.0% |
|  | *Regional* | 8 |  | 0.0% |  | 0.0% | 8 | 100.0% |
|  | *Distal* | 8 |  | 0.0% |  | 0.0% | 8 | 100.0% |
| 15 | **Asymptomatic** | **160** | **160** | **100%** |  |  |  |  |
|  | **Cancer** | **32** | **2** | 6.3% | **1** | 3.1% | **29** | 90.6% |
|  | *Local* | 16 | 2 | 12.5% | 1 | 6.3% | 13 | 81.3% |
|  | *Regional* | 8 |  | 0.0% |  | 0.0% | 8 | 100.0% |
|  | *Distal* | 8 |  | 0.0% |  | 0.0% | 8 | 100.0% |
| 16 | **Asymptomatic** | **160** | **160** | **100%** |  |  |  |  |
|  | **Cancer** | **32** | **1** | 3.1% | **2** | 6.3% | **29** | 90.6% |
|  | *Local* | 16 | 1 | 6.3% | 1 | 6.3% | 14 | 87.5% |
|  | *Regional* | 8 |  | 0.0% | 1 | 12.5% | 7 | 87.5% |
|  | *Distal* | 8 |  | 0.0% |  | 0.0% | 8 | 100.0% |
| 17 | **Asymptomatic** | **160** | **160** | **100%** |  |  |  |  |
|  | **Cancer** | **32** | **1** | 3.1% | **1** | 3.1% | **30** | 93.8% |
|  | *Local* | 16 | 1 | 6.3% |  | 0.0% | 15 | 93.8% |
|  | *Regional* | 8 |  | 0.0% | 1 | 12.5% | 7 | 87.5% |
|  | *Distal* | 8 |  | 0.0% |  | 0.0% | 8 | 100.0% |
| 18 | **Asymptomatic** | **160** | **160** | **100%** |  |  |  |  |
|  | **Cancer** | **32** | **1** | 3.1% | **1** | 3.1% | **30** | 93.8% |
|  | *Local* | 16 | 1 | 6.3% |  | 0.0% | 15 | 93.8% |
|  | *Regional* | 8 |  | 0.0% | 1 | 12.5% | 7 | 87.5% |
|  | *Distal* | 8 |  | 0.0% |  | 0.0% | 8 | 100.0% |
| 19 | **Asymptomatic** | **160** | **160** | **100%** |  |  |  |  |
|  | **Cancer** | **32** | **0** | 0.0% | **1** | 3.1% | **31** | 96.9% |
|  | *Local* | 16 |  | 0.0% |  | 0.0% | 16 | 100.0% |
|  | *Regional* | 8 |  | 0.0% | 1 | 12.5% | 7 | 87.5% |
|  | *Distal* | 8 |  | 0.0% |  | 0.0% | 8 | 100.0% |
| 20 | **Asymptomatic** | **160** | **160** | **100%** |  |  |  |  |
|  | **Cancer** | **32** | **0** | 0.0% | **1** | 3.1% | **31** | 96.9% |
|  | *Local* | 16 |  | 0.0% |  | 0.0% | 16 | 100.0% |
|  | *Regional* | 8 |  | 0.0% | 1 | 12.5% | 7 | 87.5% |
|  | *Distal* | 8 |  | 0.0% |  | 0.0% | 8 | 100.0% |

**Supplementary Table S9. Prospective Study Findings.**

| ***Sample*** | ***Samples*** | ***Negative*** | | ***Equivocal*** | | ***Positive*** | |
| --- | --- | --- | --- | --- | --- | --- | --- |
| **Benign** | **142** | **142** | **100.0%** | **-** | **-** | **-** | **-** |
| **Cancer** | **68** | **6** | 9.3% | **6** | 9.3% | **56** | 82.4% |
| *Local* | 20 | 5 | 25.0% | 2 | 10.0% | 13 | 65.0% |
| *Regional* | 7 | 1 | 14.3% | 1 | 14.3% | 5 | 71.4% |
| *Distal* | 38 | - | - | 3 | 7.9% | 35 | 92.1% |
| *Unknown* | 3 | - | - | - | - | 3 | 100.0% |

**Supplementary Table S10. CTC detection based on PSA and Gleason Score.**

**A. Overall**

|  | **PSA < 10** | **PSA 10 - 20** | **PSA > 20** | **PSA N.A.** | **Overall** |
| --- | --- | --- | --- | --- | --- |
| **GGG 1** | P: 1 (33.3%) E: - N: 2 | P: - E: 1 (100%) N: - | P: 2 (66.6%) E: - N: 1 | P: 1 (100%) E: - N: - | P: 4 (50%) E: 1 (12.5%) N: 3 |
| **GGG 2** | P: - E: - N: - | P: 2 (100%) E: - N: - | P: 2 (100%) E: - N: - | P: 2 (100%) E: - N: - | P: 6 (100%) E: - N: - |
| **GGG 3** | P: 1 (100%) E: - N: - | P: - E: - N: - | P: 4 (100%) E: - N: - | P: 1 (100%) E: - N: - | P: 6 (100%) E: - N: - |
| **GGG 4** | P: 2 (66.6%) E: - N: 1 | P: 2 (100%) E: - N: - | P: 10 (90.9%) E: - N: 1 | P: 1 (100%) E: - N: - | P: 15 (88.2%) E: - N: 2 |
| **GGG 5** | P: - E: - N: - | P: 1 (100%) E: - N: - | P: 2 (100%) E: - N: - | P: 4 (80%) E: 1 (20%) N: - | P: 7 (87.5%) E: 1 (12.5%) N: - |
| **GGG N.A.** | P: 1 (33.3%) E: 2 (66.7%) N: - | P: 2 (100%) E: - N: - | P: 11 (78.6%) E: 2 (14.3%) N: 1 | P: 4 (100%) E: - N: - | P: 18 (78.3%) E: 4 (17.4%) N: 1 |
| **Overall** | P: 5 (50%) E: 2 (20%) N: 3 | P: 7 (87.5%) E: 1 (12.5%) N: - | P: 31 (86.1%) E: 2 (5.6%) N: 3 | P: 13 (92.9%) E: 1 (7.1%) N: - | P: 56 (82.4%) E: 6 (9.3%) N: 6 |

**B. Sub-cohort where both PSA and Gleason Score were available**

|  | **PSA < 10** | **PSA 10 - 20** | **PSA > 20** | **Overall** |
| --- | --- | --- | --- | --- |
| **GGG 1** | P: 1 (33.3%) E: - N: 2 | P: - E: 1 (100%) N: - | P: 2 (66.6%) E: - N: 1 | P: 3 (50%) E: 1 (16.7%) N: 2 |
| **GGG 2** | P: - E: - N: - | P: 2 (100%) E: - N: - | P: 2 (100%) E: - N: - | P: 4 (100%) E: - N: - |
| **GGG 3** | P: 1 (100%) E: - N: - | P: - E: - N: - | P: 4 (100%) E: - N: - | P: 5 (100%) E: - N: - |
| **GGG 4** | P: 2 (66.6%) E: - N: 1 | P: 2 (100%) E: - N: - | P: 10 (90.9%) E: - N: 1 | P: 14 (87.5%) E: - N: 2 |
| **GGG 5** | P: - E: - N: - | P: 1 (100%) E: - N: - | P: 2 (100%) E: - N: - | P: 3 (100%) E: - N: - |
| **Overall** | P: 4 (66.7%) E: - N: 2 | P: 5 (83.3%) E: 1 N: - | P: 20 (90.9%) E: - N: 2 | P: 29 (85.3%) E: 1 (2.9%) N: 4 |

**Supplementary Table S11. Orthogonal Concordance Study Findings.**

| **Target (Assay)** | **Total Samples** | **Positives** | **Negatives** |
| --- | --- | --- | --- |
| AKT1_E17K | 2 | 1 | 1 |
| KRAS_G12D | 1 | 1 | 0 |
| GNAS_R201C | 1 | 1 | 0 |
| GNAS_R201H | 5 | 4 | 1 |
| TP53_R175H | 1 | 1 | 0 |
| TP53_R282W | 1 | 0 | 1 |
| PIK3CA_H1047R | 1 | 1 | 0 |
| **OVERALL** | **12** | **9 (75%)** | 3 |
